# Exploring the genetic heterogeneity of Alzheimer’s disease: Evidence for genetic subtypes

**DOI:** 10.1101/2023.05.02.23289347

**Authors:** Jeremy A. Elman, Nicholas J. Schork, Aaditya V. Rangan, the Alzheimer’s Disease Neuroimaging Initiative

## Abstract

**Background:** Alzheimer’s disease (AD) exhibits considerable phenotypic heterogeneity, suggesting the potential existence of subtypes. AD is under substantial genetic influence, thus identifying systematic variation in genetic risk may provide insights into disease origins.

**Objective:** We investigated genetic heterogeneity in AD risk through a multi-step analysis.

**Methods:** We performed principal component analysis (PCA) on AD-associated variants in the UK Biobank (AD cases=2,739, controls=5,478) to assess structured genetic heterogeneity. Subsequently, a biclustering algorithm searched for distinct disease-specific genetic signatures among subsets of cases. Replication tests were conducted using the Alzheimer’s Disease Neuroimaging Initiative (ADNI) dataset (AD cases=500, controls=470). We categorized a separate set of ADNI individuals with mild cognitive impairment (MCI; n=399) into genetic subtypes and examined cognitive, amyloid, and tau trajectories.

**Results:** PCA revealed three distinct clusters (‘constellations’) driven primarily by different correlation patterns in a region of strong LD surrounding the *MAPT* locus. Constellations contained a mixture of cases and controls, reflecting disease-relevant but not disease-specific structure. We found two disease-specific biclusters among AD cases. Pathway analysis linked bicluster-associated variants to neuron morphogenesis and outgrowth. Disease-relevant and disease-specific structure replicated in ADNI, and bicluster 2 exhibited increased CSF p-tau and cognitive decline over time.

**Conclusions:** This study unveils a hierarchical structure of AD genetic risk. Disease-relevant constellations may represent haplotype structure that does not increase risk directly but may alter the relative importance of other genetic risk factors. Biclusters may represent distinct AD genetic subtypes. This structure is replicable and relates to differential pathological accumulation and cognitive decline over time.

## BACKGROUND

Alzheimer’s disease (AD) diagnostic criteria have evolved over the years, but typically diagnosis has been characterized by predominant amnestic impairment that progressively impacts other cognitive domains and everyday functioning. However, clinical presentation is heterogeneous, and non-amnestic predominant variants of AD, often termed “atypical AD”, were specifically acknowledged in a 2011 update to AD diagnostic criteria and guidelines [1]. Pathological spread and neurodegeneration also tend to proceed in a stereotypical pattern, but mirroring the clinical diversity of AD, AD neuropathological exams and imaging studies have identified marked heterogeneity as well [2–8]. Despite this variability, AD is considered a distinct entity due to the overarching clinicopathologic characteristics observed across individuals even though the etiological basis of AD remains unclear. That is, it may have a unitary origin with diverse presentation, a highly heterogenous etiology that converges on a common disease phenotype, or it may be consistent with some intermediate scenario.

Sporadic AD is under considerable genetic influence, with an estimated heritability of 60-80% [9]. Thus, examining the genetic architecture of AD risk and the myriad ways in which combinations of variants associate with AD provides a useful foundation to understand its complex etiology. Although *APOE* represents the single largest source of genetic risk for the disease [10] recent GWAS have identified upwards of 75 different risk loci [11–15]. Considering variants that do not reach the level of genome-wide-significance but still suggest an association with AD could provide additional insight above and beyond *APOE* [16] and genome-wide significant variants. The highly oligo or polygenic nature of AD risk could reflect underlying etiological heterogeneity across individuals. The evolving definition of AD and subsequent debate over its origins further highlights the complex nature of the disease [17–19]. Given this genetic complexity yet commonality among aspects of AD clinical presentations, there is little reason to expect that phenotype-based classifications of sporadic AD (and its subtypes) will cleanly delineate homogeneous subgroups of genetic risk, providing motivation for a using “genotype-first” approach to identifying subtypes [20–22].

Identifying genetic subtypes or heterogeneity typically involves cluster analysis of some sort. There are many clustering algorithms one can exploit, and these techniques can be applied to various sources of data, including gene expression, GWAS summary statistics, and individual genotype data [23–25]. However, clustering approaches to identify genetic subtypes of disease face several difficulties (see Dahl et al. [26] for detailed discussion). First, a fundamental aspect of most clustering algorithms is that they will tend to identify clusters, even if no true clusters are present (i.e., false positive clusters). Second, a common approach is to search for clusters of genes or variants within a group of cases and consider them disease-related when this might not be the case, as it is often unknown whether these same clusters would be found in controls, which could reflect pathways or structures unrelated to disease status. The biclustering method described in Rangan et al. [27] addresses both issues. First, the biclustering method does not assume that a bicluster exists in the data, but rather tests the null hypothesis that one does *not* exist. Second, the technique searches for subsets of SNPs that express correlations within the cases that are not similarly expressed within the controls, providing evidence for disease-specificity.

Here we investigated genetic heterogeneity of Alzheimer’s disease using a combination of approaches. We first applied a principal component analysis to AD cases and controls from the UK Biobank to identify potential clusters among AD-associated SNPs that may indicate disease-relevant vulnerability across all individuals, regardless of disease status. We then applied a biclustering method to each of these likely vulnerability clusters to search for subsets of cases that harbor distinct genetic signatures that significantly increase AD risk. To validate our findings, we investigate whether the same clusters or patterns of heterogeneity also appear in an independent group of cases and controls from the Alzheimer’s Disease Neuroimaging Initiative (ADNI). Taken together, we find a hierarchical structure to an underlying heterogeneity of AD genetic risk, providing further insight into the complex etiology of the disease.

## METHODS

### Participant characteristics

We used imputed genotyping data from the UK Biobank (UKB) as the discovery dataset (**Table 1**). The UKB is a large-scale biomedical database and research resource containing genetic, lifestyle and health information from half a million UK participants [28, 29]. Data from 2,739 Alzheimer’s disease cases [30] and 5,478 age- and sex-matched controls with White British ancestry as determined by PCA [29] were included in these analyses.

**Table 1.** Sample characteristics of UK Biobank dataset.

|  | Overall | CU | AD |
| --- | --- | --- | --- |
| **n** | 8217 | 5478 | 2739 |
| **Sex, n male (%)** | 3993 (48.6) | 2662 (48.6) | 1331 (48.6) |
| **Age, years (SD)** | 64.74 (4.21) | 64.74 (4.21) | 64.74 (4.21) |
| **Education, years (SD)** | 11.95 (4.96) | 12.21 (5.01) | 11.41 (4.80) |
| **APOE-e4 alleles, n (%)** |  |  |  |
| **0** | 5082 (61.8) | 4063 (74.2) | 1019 (37.2) |
| **1** | 2647 (32.2) | 1319 (24.1) | 1328 (48.5) |
| **2** | 488 (5.9) | 96 (1.8) | 392 (14.3) |
| **APOE-e4 carriers, n (%)** | 3135 (38.2) | 1415 (25.8) | 1720 (62.8) |

CU = Cognitively unimpaired, AD = Alzheimer's disease dementia

A replication dataset was obtained from the Alzheimer’s Disease Neuroimaging Initiative (ADNI) database (adni.loni.usc.edu; **Table 2**). The ADNI was launched in 2003 as a public-private partnership, led by Principal Investigator Michael W. Weiner, MD. The primary goal of ADNI has been to test whether serial magnetic resonance imaging (MRI), positron emission tomography (PET), other biological markers, and clinical and neuropsychological assessment can be combined to measure the progression of MCI and early AD. Our biclustering analyses included genotyping data from 500 individuals with Alzheimer’s disease cases and 470 controls from the ADNI-1 (n=520), ADNI-GO/2 (n=225), and ADNI-3 (n=225). Case-control status was based on the ADNI diagnosis given at each participant’s last visit available. An additional 399 individuals (ADNI-1=166, ADNI-GO/2=175, ADNI-3=58) diagnosed with MCI at their latest visit diagnosed with MCI at their latest visit were included for examination of cognitive and biomarker change. Participants were restricted to those with primarily European ancestry (>80%) as determined by SNPweights [31].

**Table 2.** Sample characteristics of ADNI dataset. The Alzheimer’s disease (AD) and cognitively unimpaired (CU) groups were included in validation analyses of genetic heterogeneity found in the UK Biobank. The mild cognitive impairment (MCI) group was included in analyses of cognitive and biomarker trajectories.

|  | Overall | CN | Dementia | MCI |
| --- | --- | --- | --- | --- |
| **n** | 1369 | 470 | 500 | 399 |
| **Sex, n male (%)** | 765 (55.9) | 225 (47.9) | 297 (59.4) | 243 (60.9) |
| **Age, years (SD)** | 73.70 (7.11) | 72.47 (6.31) | 74.61 (7.43) | 74.00 (7.38) |
| **Education, years (SD)** | 16.01 (2.76) | 16.67 (2.44) | 15.51 (2.89) | 15.87 (2.79) |
| **APOE-e4 alleles, n (%)** |  |  |  |  |
| **0** | 750 (54.8) | 334 (71.1) | 174 (34.8) | 242 (60.7) |
| **1** | 493 (36.0) | 122 (26.0) | 245 (49.0) | 126 (31.6) |
| **2** | 126 (9.2) | 14 (3.0) | 81 (16.2) | 31 (7.8) |
| **APOE-e4 carriers, n (%)** | 619 (45.2) | 136 (28.9) | 326 (65.2) | 157 (39.3) |

### Genotyping data and quality control

Genotyping data imputed to the Haplotype Reference Consortium plus UK10K reference haplotype resource were downloaded from the UKB database along with genetic principal components (hereafter referred to as “genome-wide” principal components). Sample QC information provided by the UKB were used to apply exclusionary criteria. Individuals with excess relatedness were removed. This was defined by the UK Biobank as samples with more than 10 putative third-degree relatives (KING coefficient between 0.0442 and 0.0884). Imputed data used in these analyses included only samples with <10% missingness and biallelic SNPs with >1% minor allele frequency.

Individuals in the ADNI cohorts were genotyped using the following chips: Illumina Human610-Quad BeadChip (ADNI-1), Illumina HumanOmniExpress BeadChip (ADNI-GO/2), and Illumina Infinium Global Screening Array v2 (ADNI-3). Genetic principal components (hereafter referred to as “genome-wide” principal components) were calculated from linkage disequilibrium (LD)-pruned variants in combination with 1000 Genomes data [32] for use as covariates in later analyses. Following standard genotyping QC, imputation was performed on the Michigan Imputation Server (https://imputationserver.sph.umich.edu/) [33] using the 1000 Genomes phase 3 EUR reference panel. Imputed data from all phases were then merged. Imputed data used in this analysis included only samples with <10% missingness and biallelic SNPs with >1% minor allele frequency.

Note that, at this stage we retain all the SNPs for our primary analysis, regardless of the linkage disequilibrium (LD) relationships that might exist between them. The reason we retain all the SNPs is that our heterogeneity analysis (described below) will specifically search for combinations of SNPs that preferentially exhibit correlations across the case-subjects in contrast to the controls. These correlations drive the heterogeneous structures we are trying to find and can themselves be thought of as a form of LD that interacts with the disease. Thus, to carry out our heterogeneity analysis we retain all the relevant SNPs (regardless of LD), and later correct for ‘population-wide’ LD (e.g., SNP-correlations that are not disease-specific, but putatively ancestry-related) within our biclustering analysis below.

### Assessing high-level structure with principal components analysis

We restricted analyses to 486,823 variants in the UKB data that were associated with AD with an uncorrected p-value of <0.05 from the AD GWAS by Kunkle et al. [12] to accommodate potential association signals beyond those exhibiting genome-wide significance. After applying MAF (>1%) and genotype missingness filters (<10%), a principal components analysis (PCA) was applied to the allele combinations (dummy-coded according to the 3 possible allele combinations for each) of these AD-associated variants across all UKB cases and controls to assess the presence of high-level structure in the data (i.e., a non-Gaussian distribution). For more details on the choice of allele-coding, see Supplemental Methods section *Rationale for allele-coding.* The PCA was implemented in C using the normalized power iteration method calculated to a relative error of 1e-6, or single-digit precision. Individuals were assigned to clusters, hereafter referred to as “constellations” to disambiguate from the term “bicluster” used in subsequent steps, by applying the ISO-SPLIT algorithm [34] implemented in MATLAB (https://github.com/flatironinstitute/isosplit5) to participant loadings on the first PC.

Classification was based only on the first PC because it most robustly separated constellations across a range of p-value thresholds. Although the sample was restricted to White British participants, it is possible that these loadings reflect remaining ancestry-related population substructure. We therefore determined whether the resulting structure was specific to the set of AD-associated variants in several ways. First, PCA was applied to sets of variants restricted to p-value thresholds ranging from p<0.05 to p<1.0 in increments of 0.05. Loadings on the first two PCs were plotted at each increment, with individuals labelled according to cluster membership at the p<0.05 threshold to visually assess stability of clustering. Second, to determine whether the resulting structure was simply a function of the number of variants analyzed, a PCA was applied to a randomly chosen set of variants with the same size as those included in the analysis restricted to variants with p<0.05. Third, we examined loadings of allele combinations on the primary PC ordered by genomic position as suggested by Privé et al. [35] to assess whether it was driven by regions of high population-wide LD.

### Bicluster analysis of disease-specific structure

Heterogeneity exhibited at the top level of the constellations strongly suggests that there may be other heterogeneous substructures contained within each. This possibility seems all the more likely due to the non-Gaussian distribution of subjects within each constellation, and that the distribution of cases and controls appears to be in different directions across constellations.

To search for biclusters we use the half-loop method described in Rangan et al. [36]. This biclustering strategy (along with similar spectral-biclustering strategies) works best when the combined population of cases and controls is relatively homogenous. Given the clear structure evident from the PCA and the differential distribution of cases relative to controls across constellations, we therefore searched for disease-specific heterogeneity within each constellation separately.

Our biclustering strategy involves an iterative process which starts with all the participants (in this case, all participants in a given constellation) and SNPs, and then sequentially removes AD cases and allele-combinations from consideration. The Supplementary Methods contains a detailed description of this procedure but is summarized here. Briefly, we can measure the fraction of allele combinations that are shared between cases and subtract from this the fraction that are shared with controls to obtain a disease-related signal strength of remaining AD cases and controls. This difference can be thought of as a measure of disease-specific LD within the remaining AD-cases. The subtraction of the control-signal above is a form of “control-correction”. This control-correction is a critical step because it reduces the influence of structure present in both cases and controls on clustering. For example, disease non-specific LD or technical artifacts that are present in the full sample will be controlled for by this step. At each iteration the cases and allele-combinations with the smallest contributions to this signal are removed and the process repeats as described above. Recording this value at each step produces a ‘trace’ which indicates the disease-related signal-strength associated with the remaining AD cases and variants at that iteration.

In pursuing this process, we controlled for the first two principal-components extracted from a GRM of all individuals in our sample (and without thresholding for AD-associated SNPs) to control for ancestry. When calculating these genome-wide principal-components we did correct for LD, as our goal was to estimate the principal-components across the whole population (i.e., assuming homogeneity across subjects).

We can use the peak of the trace to delineate the membership of the dominant bicluster (i.e., which AD cases and allellic-combinations contribute to the disease-specific signal). We identify the peak by finding the internal maximum, ignoring the initial and final iterations that include >95% or <5% of AD cases.

As a null-hypothesis, we assume that the disease-label (i.e., case vs control) is not associated with the genetic profile of each subject. Therefore, we randomly permuted the case- and control-labels across subjects with similar genome-wide principal components (see Rangan et al. [27] for details) and re-calculated the traces as described above. Here, we use 500 permutations. By comparing the original trace with the distribution of traces drawn from the null-hypothesis, we can assign a p-value to the observed trace at each iteration. The null distribution will retain any structure that is uncorrelated with disease label, so provides a second type of correction against identifying clusters driven by non-disease-related LD. In this case, we only assess iterations that include >5% of the cases (i.e., the final iterations with very few cases and/or variants are ignored). To determine whether we have found a statistically significant bicluster within the original data, termed the ‘dominant’ bicluster, we can examine the p-value of the highest peak (p_max_) and the average p-value across iterations (p_avg_). Depending on the structure of the bicluster, the various p-values may be quite different, but each may be a useful metric for identifying significant biclusters. For example, if the original trace has one or more clear peaks, then there are statistically robust ‘cutpoints’ which can be used to delineate bicluster membership and p_max_ is likely to be very small. On the other hand, if the original trace has a very broad peak or a long plateau, then the bicluster is quite ‘fuzzy’, corresponding to a continuum of membership. In this case, we might expect p_avg_ to be small, but p_max_ may be relatively large. If the dominant bicluster within a data-set is statistically significant, we can extract it and then search for a secondary bicluster. This is done by scrambling the entries of the submatrix associated with the bicluster (i.e., entries corresponding to the participants and allele-combinations that were retained in the bicluster) and running the search algorithm again [27, 37].

### Gene set enrichment analysis of bicluster-associated variants

We conducted a GWAS across AD-associated SNPs comparing bicluster cases with controls belonging to the same disease-relevant constellation in which the bicluster was found (e.g., bicluster 1 cases versus controls for constellation 1) using PLINK2 [38]. Analyses were adjusted for the first two genome-wide principal components. As with the analysis of constellations, variants positively associated with each bicluster (i.e., positive regression coefficient and uncorrected p<0.05) were mapped to genes using the *g:Profiler* package in R [39]. A gene set enrichment analysis was then applied to annotated genes from each bicluster using the *clusterProfiler* package in R [40, 41]. The analysis was restricted to gene sets from the Gene Ontology resource [42, 43] containing between 10 and 1,000 genes. The Cytoscape app EnrichmentMap v3.3 [44] was used to visualize gene sets significantly enriched (FDR corrected p<0.05) in each bicluster. We constructed a network in which nodes represented gene sets significantly associated with each bicluster (gene sets can be associated with either or both).

Edges were defined by the proportion of overlapping genes between gene sets (using a minimum overlap threshold of 0.5). Next, the AutoAnnotate app (http://baderlab.org/Software/AutoAnnotate) was used to cluster and annotate nodes based on degree of overlapping genes using the MCL Cluster algorithm.

### Validation of genetic heterogeneity in ADNI

We next investigated whether the disease-relevant and disease-specific heterogeneity found in the UKB would generalize to Alzheimer’s disease cases and controls from the ADNI (see Supplementary Methods section *Replication of disease-relevant constellations in ADNI* for full details). First, we determined the subset of AD-associated SNPs common to both the UKB and ADNI datasets. Mirroring the original analysis, we applied a PCA to all cases and controls across this set of intersecting SNPs. ADNI participants were projected into the same principal components space using the allele combination loadings defined in UKB data, allowing us to assess overlap between datasets. ADNI participants were assigned to the nearest constellation based on participant loadings on the first principal component.

After participants were classified into constellations, we assessed replication of the dominant biclusters separately in the constellations in which they were found (see Supplementary Methods section *Replication of disease-specific biclusters in ADNI* for full details). Thus, we tested replication of bicluster 1 only among individuals belonging to constellation 1, and tested replication of bicluster 2 only among individuals belonging to constellation 2. For a given bicluster, we calculated the first 2 dominant SNP-wise principal components (using only SNPs in common to both datasets) among the UKB cases belonging to the bicluster. The resulting SNP loadings were used to project UKB participants in the constellation (including controls, bicluster cases, and non-bicluster cases). Similarly, we calculated projections for each of the ADNI participants belonging to a given constellation using these UKB bicluster-defined loadings. If the bicluster structure is present in the ADNI data, then the distribution of cases and controls in this space should be similar between datasets. That is, for a given participant in the testing set, participants from the training set located nearby should tend to have the same label.

We can choose what fraction of the training set participants to compare, denoted as *f*, and for a given choice of *f* we calculated the proportion of nearest neighbors in the training set that have the same label (i.e., either case or control) as each of the individuals in the test set. We assessed the quality of each choice of *f* using a permutation test in which the average “label match” across participants in the test set is compared to a null distribution constructed from label-shuffled data. We ran 500 permutations for these analyses. We calculate the average z-score across the range of *f* in (*f*_lo_, *f*_hi_), with *f*_lo_ corresponding to the lower end of the 95% confidence-interval for affine-point-matching (i.e., *f*_lo_∼ 6%) and *f*_hi_at 50%. This average is calculated using a normalizing factor to correct for heteroskedasticity [45]. The variance determining the normalizing factor is calculated from the analogous *Z*-scores obtained after alignment of the projections onto principal-components calculated from randomly selected biclusters (i.e., subsets of cases and allele-combinations) of the same size as the bicluster of interest. We calculated a global empirical p-value by comparing the average *Z*-score of the observed data across the range of parameter choices of *f* in (*f*_lo_, *f*_hi_) to the null distribution across the same range. Given that a particular bicluster is globally significant, values of *f* corresponding to high *Z*-scores indicate reasonable values of *f* to use when labelling new data, as described below.

### Association of bicluster groups with cognitive and biomarker trajectories

The procedures described above were repeated on the full set of ADNI genotyping data, this time including individuals with MCI in addition to AD cases and controls. This included applying the previously described genotyping filters, projecting ADNI data onto PC space defined by the UKB data and assigning to the nearest constellation, and then projecting individuals belonging to constellations 1 and 2 using the bicluster-defined loadings from biclusters 1 and 2, respectively. The number of nearest neighbors with each label (i.e., bicluster case, non-bicluster case, and controls) was then recorded for each ADNI participant. The fraction of training set participants used as nearest neighbors was determined by the fraction with the highest *Z*-score in the validation step described above. Each ADNI participant was assigned a soft label, calculated as the proportion of nearest neighbors in the UKB training set that were bicluster cases. To assign individuals to biclusters 1 and 2, k-means clustering with k=2 was applied to the soft labels (i.e., proportions of nearest neighbors that were bicluster cases) of each ADNI participant belonging to constellations 1 and 2 separately.

Phenotypic analyses were restricted to the ADNI MCI participants who were not included in the bicluster validation analysis. We examined change of cognition, amyloid, and phosphorylated tau (p-tau) over time between individuals labelled as bicluster 1, bicluster 2, or non-bicluster. Cognition was assessed using scores on the Preclinical Alzheimer’s Cognitive Composite (PACC) [46, 47]. Amyloid was assessed with florbetapir PET data processed according to previously published methods (http://adni.loni.usc.edu/methods) [48, 49].

Specifically, we downloaded mean standardized uptake value ratios (SUVR) from a set of regions including frontal, temporal, parietal and cingulate cortices using whole cerebellum as a reference region. Cerebrospinal fluid p-tau CSF samples were collected on cohort participants and processed as previously described [50]. CSF p-tau was measured with the fully automated Elecsys immunoassay (Roche Diagnostics) by the ADNI biomarker core (University of Pennsylvania). Florbetapir PET and CSF p-tau were chosen as the measures of amyloid and p-tau because they provided the highest number of visits with relevant data across our MCI participant group.

Group differences between individuals with MCI labelled as bicluster 1, bicluster 2, or non-bicluster were assessed with linear mixed effects models using the *lme4* [51] and *lmerTest* [52] R packages. For each outcome of interest (cognition, amyloid, or p-tau), all timepoints with available data were included. An interaction between age and group was used to assess differences in the trajectories of cognitive and biomarker measures over time. A random intercept was included for participant.

## RESULTS

### Identification of disease-relevant constellations

The PCA of AD-associated variants that were retained after applying QC filters (n=446,700 SNPs; 1,340,100 allele-combinations) in the UKB data revealed three distinct clusters, or “constellations”, each containing a mixture of AD cases and controls (**Figure 1A**). The plots in **Figure 1B** indicate that this structure only emerges when restricting to AD-associated SNPs. When all variants are included in the PCA (e.g., no p-value criterion is used) there is also evidence for substructure, but it is quite different than what is observed when a threshold of p<0.05 is used, with individuals from each of the constellations being highly interspersed. The separation of constellations along the dominant component emerges around p<0.25, and the constellations begin to further separate along the second PC at p<0.05. Results of a PCA applied to a random set of SNPs of the same size as were included at the p<0.05 level again found that individuals from each constellation were interspersed, and the overall structure closely mirrored that seen when all SNPs were included (**Supplementary** Figure 1). Three similarly distinct clusters emerge if a PCA is applied to additively-coded data as opposed to allele-coding (**Supplemental Figure 2**).

**Figure 1.**
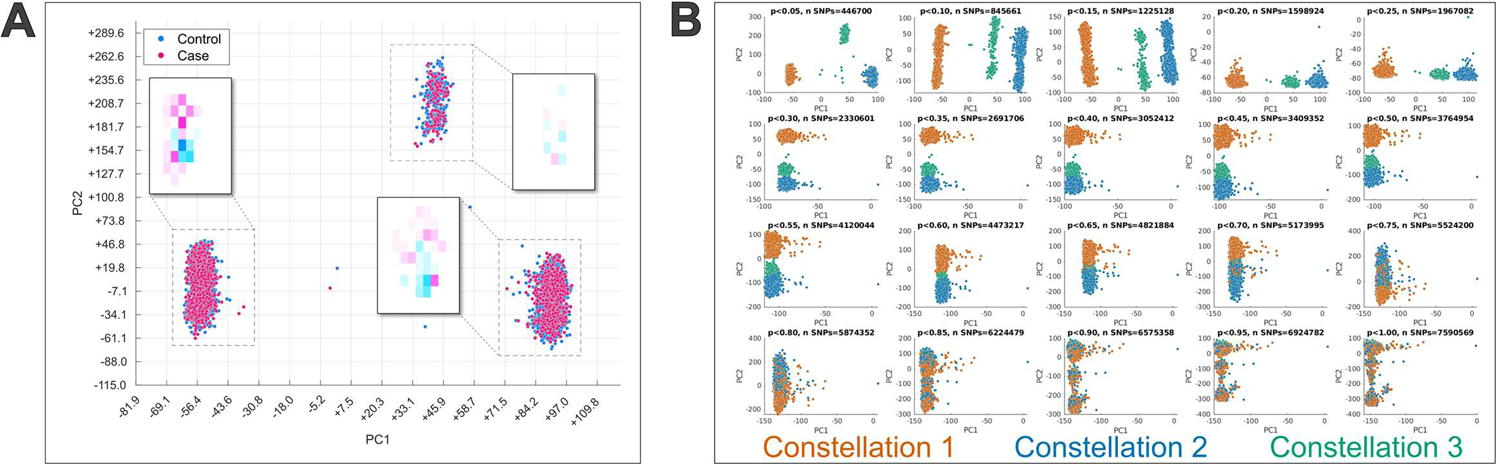
Principal component analysis of UKB data restricted to Alzheimer’s disease-associated variants reveals three constellations. **A)** Principal component analysis was applied to allele combinations of UK Biobank cases and controls restricted to variants with a p-value<0.05 in the Kunkle et al. Alzheimer’s GWAS [80]. The scatter plot displays participant loading on the first two principal components (PC1 and PC2) and colored by Alzheimer’s disease case-control status. Three distinct clusters, or constellations, are clearly present and each contains a mix of cases and controls. Heatmaps displaying the density of cases and controls in each constellation are also shown to demonstrate that, despite substantial overlap between the groups, there is some offset in the distributions. However, the directional bias is not consistent across constellations. **B)** Principal component analysis results when variants were restricted across a range of p-value thresholds from the Kunkle et al. Alzheimer’s GWAS [80]. The scatter plots are colored by constellation labels defined at the p<0.05 threshold. Participants from all three constellations are highly mixed when all variants (p<1.0) are included. The constellation structure begins to emerge along the first principal component at p<0.25, and further separate along the second principal component at p<0.05.

Plotting the SNP-wise loadings of PC1 revealed strong contributions from variants in the region of chromosome 17q21 (**Figure 2**). This is a known region of extended LD with complex genomic architecture, including a 900-kb inversion polymorphism surrounding the *MAPT* gene that defines two haplotyes, H1 and H2, with H1 containing multiple sub-haplotypes [53, 54].

**Figure 2.**
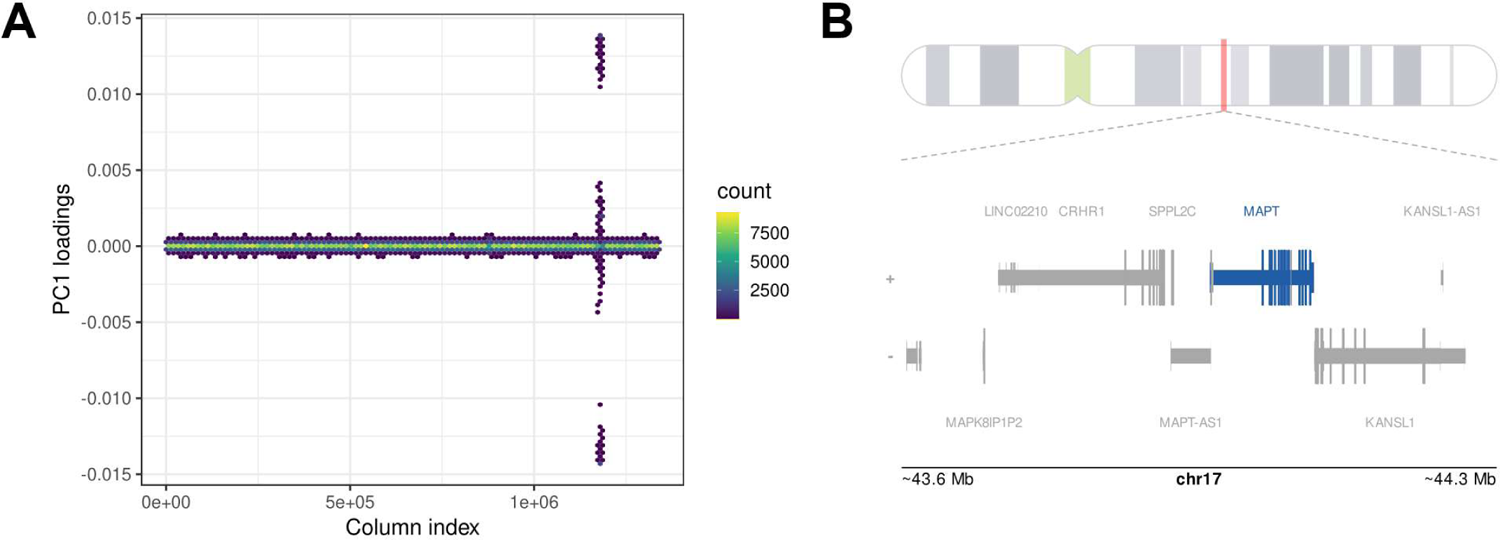
**A)** Hex bin plot of loadings across allele combinations on first principal component from the PCA on UKB cases and controls restricted to variants with a p-value<0.05 in the Kunkle et al. Alzheimer’s GWAS [12]. Allele combinations are ordered by chromosome and base position across the x-axis. Color represents density of data points that fall within a given hex. **B)** The location of peak loadings on PC1 is shown in greater detail along with gene annotations from the UCSC database. The peak loadings occur in the region of 17q21.31, overlapping with a known region of extended LD surrounding the *MAPT* locus.

Examining the genotypes of each constellation at the H1/H2 tagging SNP rs8070723, there is a strong (but not perfect) correspondence between constellations and haplotypes. Constellation 1 is largely homozygous for the H1-associated allele, constellation 3 is largely homozygous for the H2-associated allele, and constellation 2 is heterozygous (**Supplemental Table 2**). Thus, while variation from across the genome contributes to this constellation structure, it is driven primarily by several correlation patterns within this LD block, likely corresponding to H1/H2 haplotype status.

Taken together, these findings suggest that the constellations are *disease-relevant* because they only emerge as dominant modes of variation when restricting consideration to AD-associated variants, and because the distribution of cases and controls falls along different directions across constellations. However, they are not *disease-specific* because constellations contain both cases and controls, suggesting cluster membership alone does not discriminate between high and low disease risk. The three constellations were not significantly different on demographic characteristics, including sex, age, education, percent with AD dementia, or number of APOE-e4 alleles (**Supplementary Table 1**).

### Identification of disease-specific biclusters

The stark heterogeneity exhibited at the level of the disease-relevant constellations strongly suggests that there may be other heterogeneous substructures contained within each. This possibility seems all the more likely due to the non-Gaussian distribution of subjects within each constellation. Additionally, the distribution of cases and controls appears to be different across constellations. Examining **Figure 1A**, it appears that the bias or shift in cases relative to controls does not fall in the same direction across constellations, thus requiring that the search for biclusters be carried out separately for each constellation. The results of our bicluster searches are shown in **Figure 3**. Each subplot corresponds to the search in a different constellation. Within each subplot the red curve corresponds to the signal-strength of the dominant bicluster within the data, referred to as a ‘trace’ in the Methods section. The black curves indicate the distribution of traces drawn from the null-hypothesis via a permutation-test. Generally speaking, a red trace that has either a high peak or a high average (relative to the distribution of black traces) indicates a statistically significant bicluster. For this data set we detected a statistically significant bicluster in constellation 1 (termed “bicluster 1”; p_avg_=0.002, p_max_=0.008) and constellation 2 (termed bicluster 2; p_avg_=0.028, p_max_=0.244), but not in constellation 3 (p_avg_=0.484, p_max_=0.102), which is the smallest of the 3 constellations. The shape of the traces conveys additional information about the structure of the biclusters. For example, the sharp peaks defining bicluster 1 in **Figure 3A** indicate clear cut points at which membership can be clearly delineated (i.e., a disease subtype). In contrast, the broader plateau of the trace in **Figure 3B** reflects a bicluster with “fuzzy” boundaries whereby there is a smoother continuum of membership in the bicluster. This is also reflected in the different pattern of p-values between biclusters (i.e., whether the peak or average of the trace tends to be more significant). Thus, examining the traces informs us whether disease heterogeneity takes the form of distinct subtypes, a continuous spectrum of risk, or some intermediate structure. After delineating and removing the dominant bicluster within constellations 1 and 2 (see Methods) we searched once again for any additional biclusters but did not find a second bicluster that was statistically significant in either constellation (**Supplementary** Figure 3). However, when searching for these secondary biclusters, the clearly non-Gaussian distribution of traces drawn from the label-shuffled null distribution (visualized as clumped strands among the black traces) suggests there is residual heterogeneity, but we are not powered to identify it in the current sample. The bicluster participants were not significantly different on demographic characteristics, including sex, age, education, or number of APOE-e4 alleles (**Supplementary Table 1**). **Supplementary** Figure 4 presents a plot of the constellations shown in **Figure 1** with bicluster cases highlighted.

**Figure 3.**
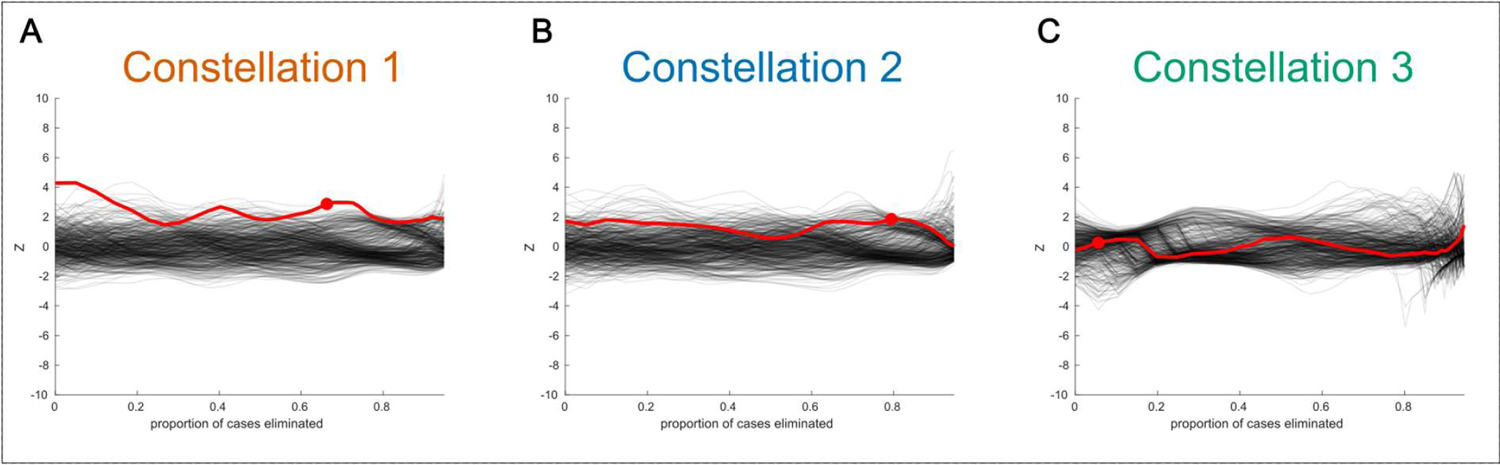
Bicluster traces of observed data versus label-shuffled data. The disease-related signal-strength associated with the remaining UKB Alzheimer’s cases relative to controls is plotted on the y-axis. At each iteration, allele combinations and cases that contribute least to this difference are removed. The proportion of remaining cases is shown on the x-axis. The red trace represents the original data and black traces represent label-shuffled data, corresponding to a null distribution. A red dot indicates the iteration with the maximum separation between cases and controls (ignoring signal in the first iterations), and is used to define the bicluster. The sharper peak of constellation 1 indicates that this bicluster has more distinct boundaries, whereas the bicluster in constellation 2 has “fuzzier” boundaries as indicated by the broad yet lower peak.

### Gene set enrichment results of disease-specific biclusters

Bicluster 1 was enriched for a number of gene sets, including those related to receptor activity, calcium and sodium ion transport, dendritic structure, GTPase activity, and regulation of the MAPK cascade. Bicluster 2 was enriched for gene sets related to the MHC protein complex, regulation of cell size, lipid transport, and tyrosine kinase activity. Both biclusters showed enrichment for cell-projection morphogenesis, and synaptic transmission. **Figure 4** shows network plots of gene sets enriched in each bicluster. The gene sets are grouped by proportion of overlapping genes and labelled with dominant terms to illustrate the shared and unique functions associated with variants in each bicluster.

**Figure 4.**
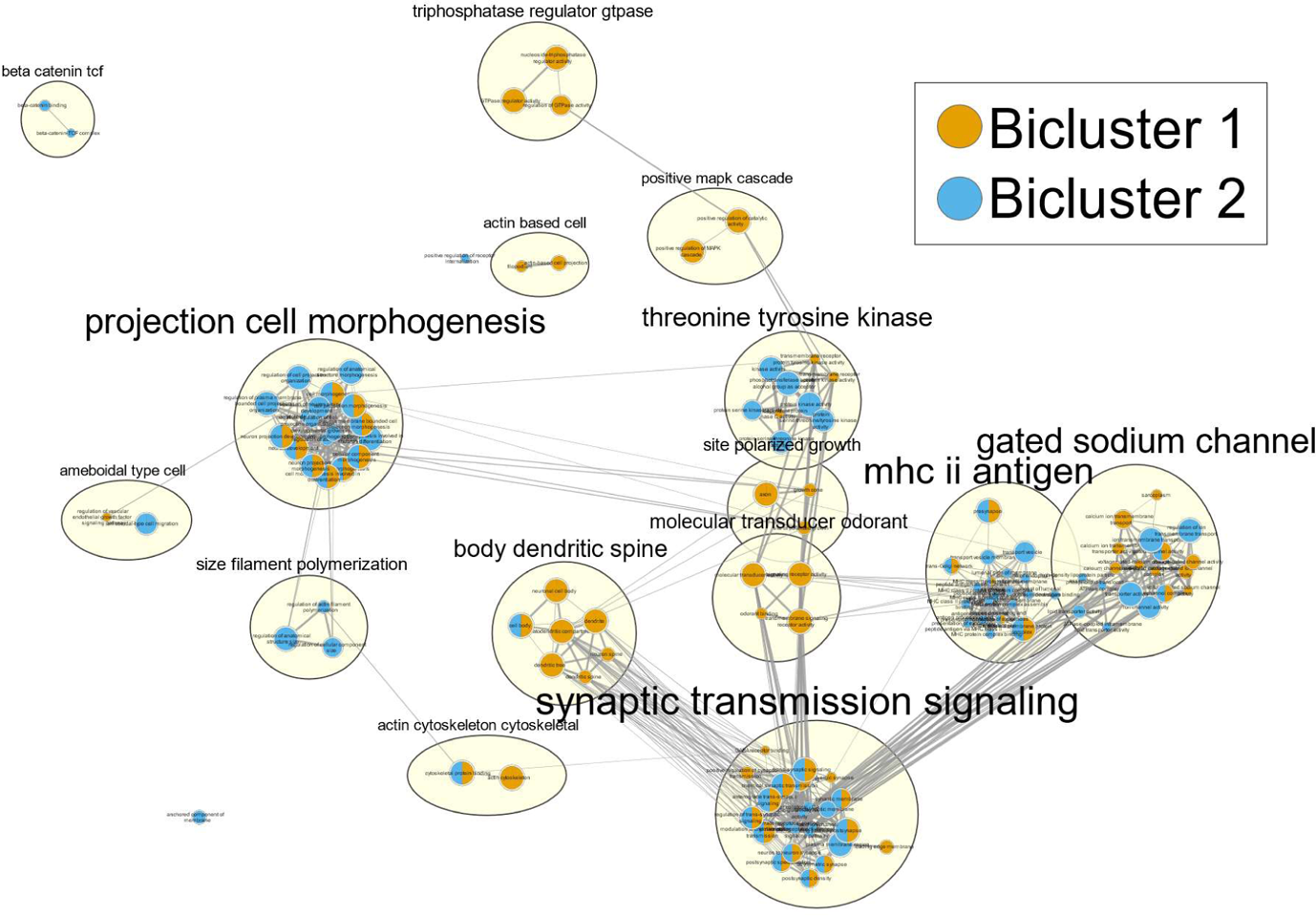
Network plot of gene sets enriched among bicluster genes. Network visualization of gene set enrichment results for biclusters 1 and 2. First, separate GWAS compared individuals in each bicluster to all controls from the same constellation in which the bicluster was found (e.g., bicluster 1 cases versus all controls from constellation 1). SNPs that were nominally associated with a given bicluster (i.e., positive regression coefficient and p<0.05) were mapped to genes based on position. Over-representation analysis of gene lists for each bicluster was used to identify gene sets associated with each bicluster. Nodes represent significantly enriched gene sets (p_FDR_<0.05) with color indicating the bicluster they are associated with. Edges represent the overlap in genes belonging to gene sets using a threshold of 0.5. Gene sets were clustered based on overlaps and automatically annotated based on the descriptions of each gene set cluster.

Although many of the enriched gene sets identified in this analysis relate to basic functions and thus do not appear to be AD-specific, it should be noted that the variants considered in these analyses were already selected for their association with AD. That is, the gene sets shown in **Figure 4** are those which are enriched in bicluster variants relative to the other AD-associated variants, rather than with respect to all other variants, including AD-nonspecific variants. Pathways that are typically found to be enriched for AD-associated variants constitute a background signal that is likely shared among many of the cases, including those that are not part of either bicluster.

There is also a degree of overlap in the gene sets enriched for each bicluster. However, the proportion of overlap seen at the level of gene sets is reduced when looking at the overlap among constituent genes or SNPs (**Supplementary** Figure 5). This indicates that the variants associated with each bicluster may have impacts that converge on similar downstream pathways, but the specific perturbations encompassed by each may not be the same.

### Disease-relevant constellations are evident in ADNI data

Using only SNPs common to both datasets, we re-calculated the top principal components of AD-associated variants in the UKB data. These loadings were used to project both the UKB and ADNI data into the same PC space. As seen in **Figure 5**, the grouping of UKB participants into 3 constellations was almost perfectly mirrored in the ADNI data, albeit with a slight shift in positioning. As in the UKB data, each constellation contained a mixture of AD cases and controls, reflecting disease-relevant but not disease-specific clustering.

**Figure 5.**
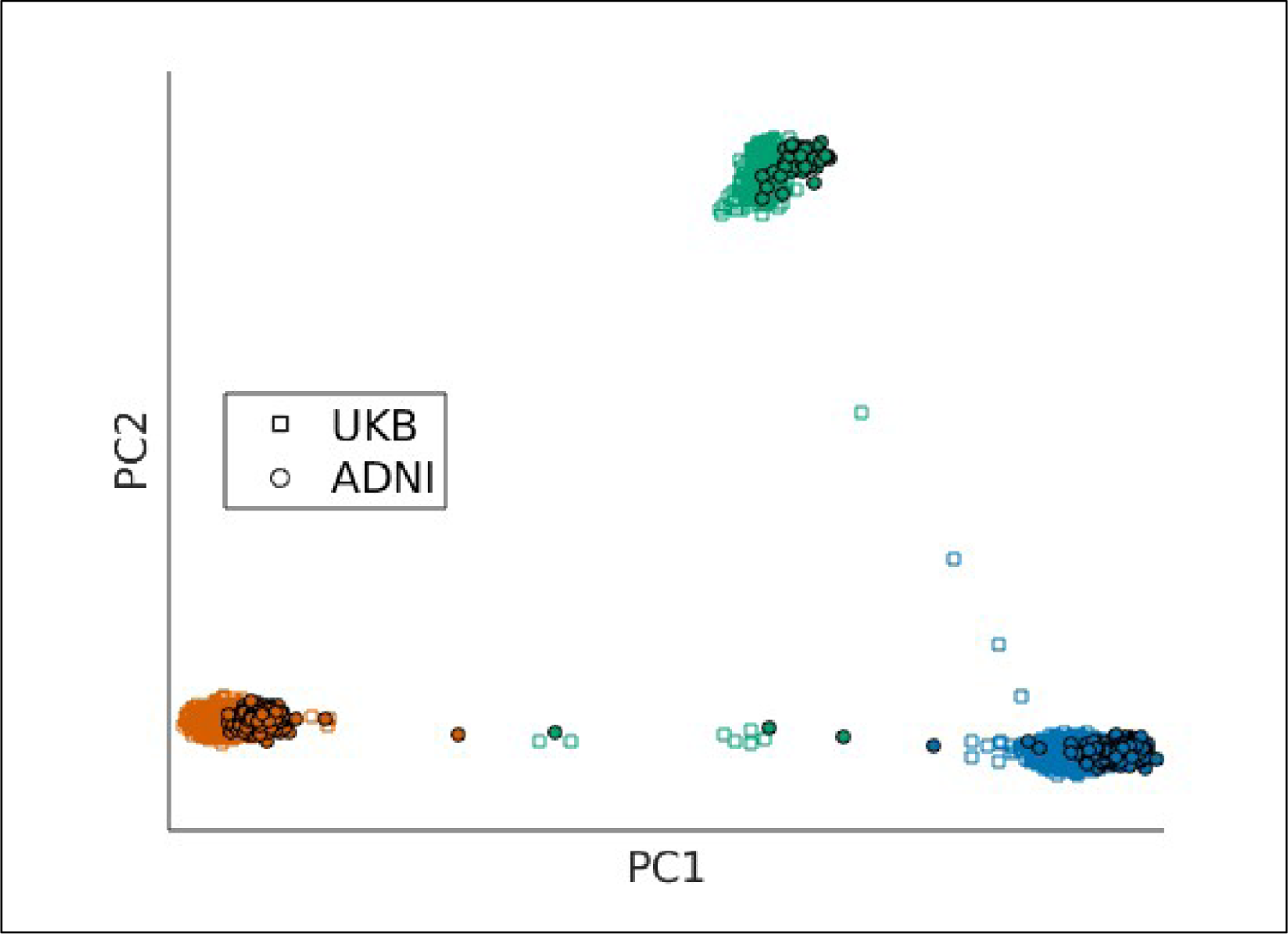
ADNI data projected along principal components defined in UKB data replicate distribution of disease-relevant constellations. Principal components were recalculated in UKB data using only AD-associated variants common to both datasets. Participants from both datasets were then plotted by participants loadings on the first two principal components. Colors represent constellations, UKB participants are plotted with squares, and ADNI participants are plotted with circles.

### Disease-specific biclusters replicate in ADNI

Overall, we found evidence for significant replication of bicluster 1 in the ADNI data (global p=0.006), and weaker yet significant replication of bicluster 2 (global p=0.048). This pattern is consistent with the initial bicluster search, in which bicluster 1 seemed to have more distinct boundaries whereas the boundaries of bicluster 2 were more diffuse. The best label similarity ranking in comparison to the null distribution was achieved using 31.6% of the sample as nearest neighbors for bicluster 1 and using 27.3% of the sample as nearest neighbors for bicluster 2. **Figure 6** displays the labeling accuracy across a range of nearest neighbors. Note that the shape of these curves can be influenced by the structure of the bicluster. For example, a bicluster with clearly delineated boundaries may be less sensitive to altering the nearest neighbor parameter compared to one with blurrier boundaries. Consistent with this, we see high accuracy of label matching in bicluster 1 across a large range of nearest neighbor fractions, ranging from about 17% up to our maximum threshold of 50%. In contrast, the accuracy of label matching bicluster 2 is more sensitive to this parameter choice, with high accuracy using nearest neighbor fractions from about 26% to 36%. Importantly, these plots provide a range of sensible values one can use to label new data.

**Figure 6.**
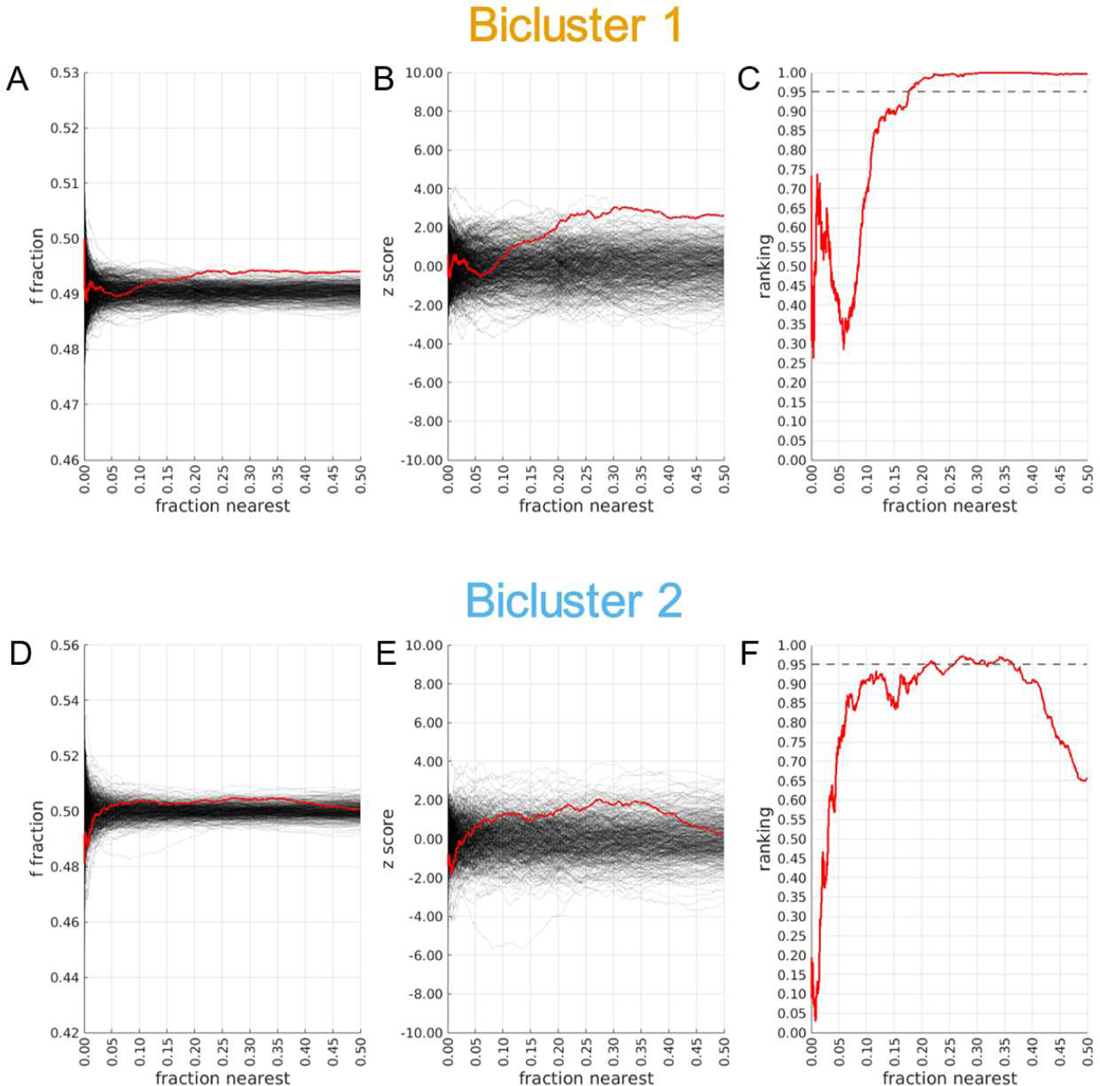
Replication of disease-specific biclusters in ADNI data. The similarity between case-control labels of individuals in the test set (ADNI) and the most frequent label among nearest neighbors in the training set (UKB) was used to assess replication of biclusters. Only individuals belonging to the constellation in which the given bicluster was found were considered. The fraction of individuals from the training set (i.e., UKB) considered as nearest neighbors is plotted along the x-axis. The y-axis in Panels (**A**) and (**D**) shows the average fraction of nearest neighbors with a matching label. The red line in Panels (**A**) and (**D**) shows values from the original data, while the black lines show values from label-shuffled trials drawn from the null-distribution. A trial-wise mean and variance can be defined from the null distribution to normalize values, with the associated z-scores shown in Panels (**B**) and (**E**). A rank-normalization of the scores compared to the null distribution is shown in Panels (**C**) and (**F**).

### Cognitive and biomarker trajectories across bicluster groups in ADNI MCI participants

We found evidence for differential cognitive and biomarker trajectories across bicluster groups in a separate set of ADNI individuals diagnosed with MCI. Individuals assigned to bicluster 2 demonstrated significantly greater decline on the PACC compared to the non-bicluster (β=-0.40, t-value=-2.29, p=0.022) and bicluster 1 groups (β=-0.60, t-value=2.92, p=0.004) (**Figure 7A**). Bicluster 1 demonstrated a somewhat greater increase in florbetapir over time compared to the non-bicluster group, but this difference was not significant (β=0.32, t-value=1.52, p=0.129) (**Figure 7B**). Bicluster 2 demonstrated a significantly greater increase of CSF p-tau over time compared to the non-bicluster group (β=0.45, t-value=2.05, p=0.041) (**Figure 7C**).

**Figure 7.**
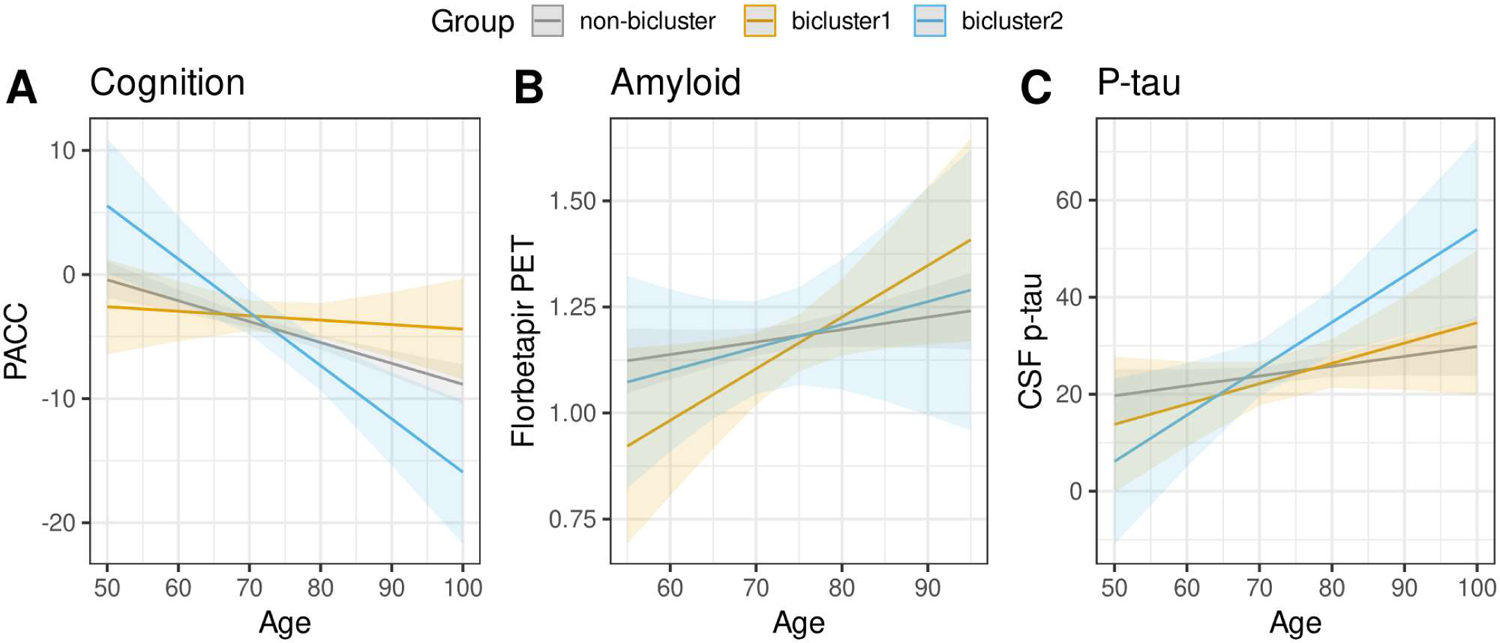
Association of bicluster groups in ADNI MCI sample. ADNI individuals diagnosed with MCI were assigned to bicluster groups (bicluster 1, bicluster 2, or non-bicluster). Differences in longitudinal cognitive and biomarker trajectories between groups were tested with age x group interactions in linear mixed effects models. Model predicted values are shown in the figure. A) Cognition was measured with the Preclinical Alzheimer’s Cognitive Composite (PACC). B) Amyloid was measures using florbetapir PET. C) Phosphorylated tau (p-tau) was measured using CSF p-tau.

## DISCUSSION

Genetic analyses suggest the etiology of AD is multifactorial [55], but the extent to which the composition of genetic influences on AD risk varies across individuals and whether it is replicable across different data sets has been unclear. We identified evidence for several subsets of individuals that exhibit different patterns of genetically-mediated vulnerability to the risk for AD. Among AD-associated SNPs, there is clear heterogeneity across individuals, and this heterogeneity seemed to follow a hierarchical structure. Importantly, this structure was observed in two independent datasets, indicating that it is not sample-specific, but rather appears to be a generalizable feature of AD genetic risk.

The first level of heterogeneity emerged as 3 clusters, which we termed “constellations”, from a PCA of the UKB data confined to variants associated with AD based on prior GWAS. Similar analyses (i.e., PCA) are often used to identify ancestry-related population structure [56], but we found that the constellations only emerged when examining AD-associated SNPs and individuals from each constellation were highly intermixed when analyzed using all SNPs or a random selection of SNPs. These results indicate that the constellations do not simply reflect ancestry-related substructure. We further found a spike in loadings on the first PC used to define the constellations within the region of chromosome 17q21.31. This is a region of strong extended LD driven by a 900-kb inversion polymorphism surrounding the *MAPT* gene that defines two haplotypes, H1 and H2, with the H1 haplotype further dividing into several sub-haplotypes [53, 54]. The *MAPT* gene codes for tau, the primary component of neurofibrillary tangles, so it is clearly relevant to AD. However, as a risk gene it is more strongly associated with primary tauopathies, and investigations of the *MAPT* locus and the H1/H2 haplotypes find inconsistent evidence for a specific association with AD risk [54, 57]. A stratified GWAS of AD by Strickland, et al. [58] found that there were several variants with haplotype-dependent associations, which may explain these inconsistencies.

It may be that our constellations at least partially reflect the H1/H2 haplotypes (along with H1 sub-haplotypes) defined by variation in this region. Using the H1/H2 haplotype-tagging SNP rs8070723, Strickland, et al. [58] found that 39% of their sample were H2 carriers (H1H2 + H2H2). Based on the genotypes of individuals across our constellations, individuals in constellations 2 and 3 (who largely have genotypes corresponding to H1H2 and H2H2 carriers, respectively) accounted for an identical 39% in our sample as well. A block of complete LD in this region and minimal recombination may further explain the surprising degree of separation between these constellations [53, 54, 59]. We note an important aspect of these constellations: they contain both cases and controls and therefore membership is not associated with increased AD risk. However, the overall distribution of cases and controls is different in each. Taken together with results from Strickland, et al. [58] showing haplotype-dependent associations, we refer to the constellation structure are “disease-relevant” in that it may alter the relative importance of other risk variants (e.g., by associating with protection or vulnerability to these other risk variants) but is not directly associated with increased AD risk.

Cases belonging to the disease-specific biclusters harbored distinct genetic signatures when compared to other AD cases and may therefore reflect “subtypes” of AD genetic risk. The variants defining the two disease-specific biclusters were enriched for both similar and unique pathways when compared to each other and the disease-relevant constellations in which they were found. It is important to re-emphasize the fact that these analyses were restricted to AD-associated variants, which are enriched for lipid processing, cholesterol transport, amyloid precursor (APP) processing and Aβ formation, tau protein binding, and immune response [12]. The bicluster-associated pathways reported here thus exist within this broader context of other AD-associated pathways. Bicluster 1 was enriched for gene sets involved in regulating the MAPK cascade, which may contribute to AD progression in several cell-type-dependent ways, such as increasing neuroinflammation, promoting neurofibrillary tangle formation, and depressing synaptic plasticity [60]. Bicluster 2 demonstrated enrichment in a cluster of gene sets related to the major histocompatibility complex (MHC), which is consistent with findings that implicate microglia-mediated immune response as a key player in AD [12, 15, 61, 62]. Both biclusters – and bicluster 1 in particular – were enriched for gene sets related to synaptic signaling and signal transduction. Similar enrichment was found among newly prioritized genes in the most recent largescale GWAS of AD [15]. We also found both biclusters were strongly enriched for a number of pathways that interact to influence the morphogenesis and outgrowth of neurons. This includes gene sets related to the cellular components themselves (e.g., cell bodies, dendritic spines, synapses, axonal projections, cystoskeletal components), but also factors that modulate their development such as polymerization, β-catenin binding, and GTPase activity [63–67]. Disruption of these processes may leave structures more vulnerable to insult resulting in, for example, greater Aβ-mediated synapse loss. APP (the precursor of Aβ) is important for regulating axonal and synaptic growth through influencing cytoskeletal remodelling [68] and tau plays a critical role in stabilizing microtubules [69, 70]. It is therefore possible that the hallmark pathologies of AD, i.e., Aβ and tau protein build-up, may exert impacts through toxic effects, but also reflect a loss of function that affects cell structure and growth. Further work is needed to comprehensively characterize the impacts of these bicluster-associated variants.

We found that both the disease-relevant constellation structure and the disease-specific biclusters replicated in the ADNI sample. There are several important differences between the ADNI and UKB cohorts. First, although analyses were restricted to individuals of European ancestry, the UKB was further restricted to a White British subset, so the ADNI data may contain relatively more ancestry-related heterogeneity. Second, the studies used different genotyping chips (and ADNI genotypes were obtained with 3 different chips across phases) and imputation panels. Third, ADNI participants were recruited with the intent of mirroring a clinical trial population focused on Alzheimer’s disease, whereas the UKB attempted to recruit a broader base of individuals to study multiple outcomes. Replicable structure across these datasets is thus unlikely to reflect dataset-specific or artifactual confounds that should differentially affect these datasets.

Supporting the relevance of the genetic structure found here, we found that a separate set of ADNI participants diagnosed with MCI assigned to different bicluster groups demonstrated differential cognitive and biomarker trajectories. Individuals with genetic signatures resembling bicluster 2 exhibited greater accumulation of p-tau and a corresponding steeper decline in cognitive performance over time. The variants associated with this genetic subtype may contribute to a more aggressive form of AD. We previously found that genetic risk affecting different biological pathways can preferentially relate to amyloid or tau accumulation [71]. Thus, it may be that the balance of pathological accumulation is shifted towards p-tau in these individuals, and p-tau has been shown to be more closely linked to subsequent neurodegeneration [72] and cognitive decline [73] compared to amyloid. The genetic heterogeneity observed here may impact not only level but distribution of pathology, which may be particularly important in the case of tau [3, 7, 74]. Heterogeneity in genetic risk may also relate to comorbid conditions that exacerbate AD progression. It is therefore clear that more work is needed to characterize the downstream impacts of genetic heterogeneity on disease outcomes.

Our findings may indicate that there are likely multiple genetically-mediated pathologies underlying AD that converge on common clinical manifestations. This is not a unique phenomenon. For example, Charcot-Marie-Tooth disease involves a common set of clinical symptoms that can arise from separate genetic origins [75]. We did find evidence for overlap in the pathways enriched across the different constellations and biclusters identified in our analysis, and this may indicate where convergence begins. However, there are a few caveats. First, receiving the same clinical diagnosis does not rule out the possibility of meaningful differences across cases. There are atypical forms of clinically-defined AD [76–79] and there is growing evidence for several biological subtypes of AD characterized by distinct patterns of pathological spread or neurodegeneration [2, 3, 5]. Further work is needed to determine whether the genetic heterogeneity identified here is associated with more subtle phenotypic differences. Second, the degree of overlap among pathways that we observed is driven by SNPs that were assigned to genes and pathways, but not all SNPs were assigned to genes.

Many AD-associated SNPs are located in non-coding regions, so the overlap of enriched gene sets ignores substantial numbers of SNPs uniquely associated with each. Differences between constellations and biclusters may arise from the functional effects of the variants themselves, including those not assigned to genes. Third, it is possible that the subsets of cases identified by the biclustering represent misdiagnosed AD (assuming the underlying disease cause is driven by partially distinct genetic factors). Identifying misdiagnoses or undiagnosed cases in a heterogenuous population is a potential use-case of the method. However, the number of cases in each bicluster would require a higher rate of misdiagnosis than one might expect, especially in ADNI where other dementias were specifically excluded.

## CONCLUSIONS

In sum, we found evidence of a hierarchical structure underlying heterogeneity in the genetic risk of AD. The disease-relevant constellation structure is driven to a large degree by an extended region of LD on chromosome 17q.21, with constellations potentially reflecting *MAPT* haplotypes. Membership in a given constellation did not directly increase risk for AD, but may alter the relative importance of other genetic risk variants for AD. On the other hand, the biclusters may be considered disease subtypes with distinct genetic signatures compared to the broader population of AD cases, which may have important implications for treatment efforts. Despite all cases presenting with a common clinical syndrome, the etiology and path taken to clinical manifestation may vary across patients. The differential cognitive and biomarker trajectories between genetic subtypes provides some evidence that this heterogeneity has consequences for downstream disease processes. Identifying subtypes of AD could facilitate precision medicine approaches that tailor treatment strategies to the individual for increased effectiveness.

## Data Availability

Data used in preparation of this article were obtained from the UK Biobank (application ID 63648) database at http://www.ukbiobank.ac.uk/ and the Alzheimer’s Disease Neuroimaging Initiative (ADNI) database at https://adni.loni.usc.edu/. These data may be obtained upon application to the respective studies and completion of data use and/or material transfer agreements. Code implementing the bicluster search is available at https://github.com/adirangan/lakcluster_matlab. Additional code used in the analysis is available upon request to the corresponding author (J.A.E.).

## Acknowledgments

We thank the anonymous reviewers whose insightful comments and suggestions helped improve the manuscript. This research has been conducted using data from UK Biobank, a major biomedical database (http://www.ukbiobank.ac.uk/). Data collection and sharing for this project was funded by the Alzheimer’s Disease Neuroimaging Initiative (ADNI) (National Institutes of Health Grant U01 AG024904) and DOD ADNI (Department of Defense award number W81XWH-12-2-0012). ADNI is funded by the National Institute on Aging, the National Institute of Biomedical Imaging and Bioengineering, and through generous contributions from the following: AbbVie, Alzheimer’s Association; Alzheimer’s Drug Discovery Foundation; Araclon Biotech; BioClinica, Inc.; Biogen; Bristol-Myers Squibb Company; CereSpir, Inc.; Cogstate; Eisai Inc.; Elan Pharmaceuticals, Inc.; Eli Lilly and Company; EuroImmun; F. Hoffmann-La Roche Ltd and its affiliated company Genentech, Inc.; Fujirebio; GE Healthcare; IXICO Ltd.; Janssen Alzheimer Immunotherapy Research & Development, LLC.; Johnson & Johnson Pharmaceutical Research & Development LLC.; Lumosity; Lundbeck; Merck & Co., Inc.; Meso Scale Diagnostics, LLC.; NeuroRx Research; Neurotrack Technologies; Novartis Pharmaceuticals Corporation; Pfizer Inc.; Piramal Imaging; Servier; Takeda Pharmaceutical Company; and Transition Therapeutics. The Canadian Institutes of Health Research is providing funds to support ADNI clinical sites in Canada. Private sector contributions are facilitated by the Foundation for the National Institutes of Health (www.fnih.org). The grantee organization is the Northern California Institute for Research and Education, and the study is coordinated by the Alzheimer’s Therapeutic Research Institute at the University of Southern California. ADNI data are disseminated by the Laboratory for Neuro Imaging at the University of Southern California.

## Funding

This work was supported by National Institute on Aging grants K01 AG063805 (JAE), UH3-AG064706 (NJS), U19 AG023122 (NJS), U19 AG065169-01A1 (NJS) and U24AG078753 (NJS).

## Conflict of interest

Jeremy Elman is an Editorial Board Member of this journal but was not involved in the peer-review process of this article nor had access to any information regarding its peer-review. All other authors have no conflict of interest to report.

## Notes

### Competing Interest Statement

The authors have declared no competing interest.

### Funding Statement

National Institute on Aging grant K01 AG063805 to JAE
National Institute on Aging grants UH3-AG064706, U19 AG023122, U19 AG065169-01A1 and U24AG078753 to NJS
Data collection and sharing for this project was funded by the Alzheimer's Disease Neuroimaging Initiative (ADNI) (National Institutes of Health Grant U01 AG024904) and DOD ADNI (Department of Defense award number W81XWH-12-2-0012).

### Author Declarations

The North West Multi-centre Research Ethics Committee (MREC) gave ethical approval for the collection and use of UK Biobank data. Procedures used in the Alzheimer's Disease Neuroimaging Initiative were approved by the Institutional Review Board of participating institutions. The University of California San Diego Human Research Protections Program gave ethical approval for this work.

### Summary of Updates

Revised main text and figures; Revised Supplemental Files

## REFERENCES

[1] McKhann GM, Knopman DS, Chertkow H, Hyman BT, Jack CR, Jr., Kawas CH, Klunk WE, Koroshetz WJ, Manly JJ, Mayeux R, Mohs RC, Morris JC, Rossor MN, Scheltens P, Carrillo MC, Thies B, Weintraub S, Phelps CH (2011) The diagnosis of dementia due to Alzheimer’s disease: recommendations from the National Institute on Aging-Alzheimer’s Association workgroups on diagnostic guidelines for Alzheimer’s disease. Alzheimers Dement 7, 263–269.

[2] Murray ME, Graff-Radford NR, Ross OA, Petersen RC, Duara R, Dickson DW (2011) Neuropathologically defined subtypes of Alzheimer’s disease with distinct clinical characteristics: a retrospective study. Lancet Neurol 10, 785–796.

[3] Vogel JW, Young AL, Oxtoby NP, Smith R, Ossenkoppele R, Strandberg OT, La Joie R, Aksman LM, Grothe MJ, Iturria-Medina Y, Alzheimer’s Disease Neuroimaging I, Pontecorvo MJ, Devous MD, Rabinovici GD, Alexander DC, Lyoo CH, Evans AC, Hansson O (2021) Four distinct trajectories of tau deposition identified in Alzheimer’s disease. Nat Med 27, 871–881.

[4] Young AL, Marinescu RV, Oxtoby NP, Bocchetta M, Yong K, Firth NC, Cash DM, Thomas DL, Dick KM, Cardoso J, van Swieten J, Borroni B, Galimberti D, Masellis M, Tartaglia MC, Rowe JB, Graff C, Tagliavini F, Frisoni GB, Laforce R, Jr., Finger E, de Mendonca A, Sorbi S, Warren JD, Crutch S, Fox NC, Ourselin S, Schott JM, Rohrer JD, Alexander DC, Genetic FTDI, Alzheimer’s Disease Neuroimaging I (2018) Uncovering the heterogeneity and temporal complexity of neurodegenerative diseases with Subtype and Stage Inference. Nat Commun 9, 4273.

[5] Ferreira D, Nordberg A, Westman E (2020) Biological subtypes of Alzheimer disease: A systematic review and meta-analysis. Neurology 94, 436–448.

[6] Ferreira D, Verhagen C, Hernandez-Cabrera JA, Cavallin L, Guo CJ, Ekman U, Muehlboeck JS, Simmons A, Barroso J, Wahlund LO, Westman E (2017) Distinct subtypes of Alzheimer’s disease based on patterns of brain atrophy: longitudinal trajectories and clinical applications. Sci Rep 7, 46263.

[7] Ossenkoppele R, Schonhaut DR, Scholl M, Lockhart SN, Ayakta N, Baker SL, O’Neil JP, Janabi M, Lazaris A, Cantwell A, Vogel J, Santos M, Miller ZA, Bettcher BM, Vossel KA, Kramer JH, Gorno-Tempini ML, Miller BL, Jagust WJ, Rabinovici GD (2016) Tau PET patterns mirror clinical and neuroanatomical variability in Alzheimer’s disease. Brain 139, 1551–1567.

[8] Tijms BM, Gobom J, Reus L, Jansen I, Hong S, Dobricic V, Kilpert F, ten Kate M, Barkhof F, Tsolaki M, Verhey FRJ, Popp J, Martinez-Lage P, Vandenberghe R, Lleó A, Molinuevo JL, Engelborghs S, Bertram L, Lovestone S, Streffer J, Vos S, Bos I, The Alzheimer’s Disease Neuroimaging I, Blennow K, Scheltens P, Teunissen CE, Zetterberg H, Visser PJ (2020) Pathophysiological subtypes of Alzheimer’s disease based on cerebrospinal fluid proteomics. Brain.

[9] Gatz M, Reynolds CA, Fratiglioni L, Johansson B, Mortimer JA, Berg S, Fiske A, Pedersen NL (2006) Role of genes and environments for explaining Alzheimer disease. Arch Gen Psychiatry 63, 168–174.

[10] Corder EH, Saunders AM, Strittmatter WJ, Schmechel DE, Gaskell PC, Small GW, Roses AD, Haines JL, Pericak-Vance MA (1993) Gene dose of apolipoprotein E type 4 allele and the risk of Alzheimer’s disease in late onset families. Science 261, 921–923.

[11] Harold D, Abraham R, Hollingworth P, Sims R, Gerrish A, Hamshere ML, Pahwa JS, Moskvina V, Dowzell K, Williams A, Jones N, Thomas C, Stretton A, Morgan AR, Lovestone S, Powell J, Proitsi P, Lupton MK, Brayne C, Rubinsztein DC, Gill M, Lawlor B, Lynch A, Morgan K, Brown KS, Passmore PA, Craig D, McGuinness B, Todd S, Holmes C, Mann D, Smith AD, Love S, Kehoe PG, Hardy J, Mead S, Fox N, Rossor M, Collinge J, Maier W, Jessen F, Schurmann B, Heun R, van den Bussche H, Heuser I, Kornhuber J, Wiltfang J, Dichgans M, Frolich L, Hampel H, Hull M, Rujescu D, Goate AM, Kauwe JS, Cruchaga C, Nowotny P, Morris JC, Mayo K, Sleegers K, Bettens K, Engelborghs S, De Deyn PP, Van Broeckhoven C, Livingston G, Bass NJ, Gurling H, McQuillin A, Gwilliam R, Deloukas P, Al-Chalabi A, Shaw CE, Tsolaki M, Singleton AB, Guerreiro R, Muhleisen TW, Nothen MM, Moebus S, Jockel KH, Klopp N, Wichmann HE, Carrasquillo MM, Pankratz VS, Younkin SG, Holmans PA, O’Donovan M, Owen MJ, Williams J (2009) Genome-wide association study identifies variants at CLU and PICALM associated with Alzheimer’s disease. Nat Genet 41, 1088–1093.

[12] Kunkle BW, Grenier-Boley B, Sims R, Bis JC, Damotte V, Naj AC, Boland A, Vronskaya M, van der Lee SJ, Amlie-Wolf A, Bellenguez C, Frizatti A, Chouraki V, Martin ER, Sleegers K, Badarinarayan N, Jakobsdottir J, Hamilton-Nelson KL, Moreno-Grau S, Olaso R, Raybould R, Chen Y, Kuzma AB, Hiltunen M, Morgan T, Ahmad S, Vardarajan BN, Epelbaum J, Hoffmann P, Boada M, Beecham GW, Garnier JG, Harold D, Fitzpatrick AL, Valladares O, Moutet ML, Gerrish A, Smith AV, Qu L, Bacq D, Denning N, Jian X, Zhao Y, Del Zompo M, Fox NC, Choi SH, Mateo I, Hughes JT, Adams HH, Malamon J, Sanchez-Garcia F, Patel Y, Brody JA, Dombroski BA, Naranjo MCD, Daniilidou M, Eiriksdottir G, Mukherjee S, Wallon D, Uphill J, Aspelund T, Cantwell LB, Garzia F, Galimberti D, Hofer E, Butkiewicz M, Fin B, Scarpini E, Sarnowski C, Bush WS, Meslage S, Kornhuber J, White CC, Song Y, Barber RC, Engelborghs S, Sordon S, Voijnovic D, Adams PM, Vandenberghe R, Mayhaus M, Cupples LA, Albert MS, De Deyn PP, Gu W, Himali JJ, Beekly D, Squassina A, Hartmann AM, Orellana A, Blacker D, Rodriguez-Rodriguez E, Lovestone S, Garcia ME, Doody RS, Munoz-Fernadez C, Sussams R, Lin H, Fairchild TJ, Benito YA, Holmes C, Karamujic-Comic H, Frosch MP, Thonberg H, Maier W, Roshchupkin G, Ghetti B, Giedraitis V, Kawalia A, Li S, Huebinger RM, Kilander L, Moebus S, Hernandez I, Kamboh MI, Brundin R, Turton J, Yang Q, Katz MJ, Concari L, Lord J, Beiser AS, Keene CD, Helisalmi S, Kloszewska I, Kukull WA, Koivisto AM, Lynch A, Tarraga L, Larson EB, Haapasalo A, Lawlor B, Mosley TH, Lipton RB, Solfrizzi V, Gill M, Longstreth WT, Jr., Montine TJ, Frisardi V, Diez-Fairen M, Rivadeneira F, Petersen RC, Deramecourt V, Alvarez I, Salani F, Ciaramella A, Boerwinkle E, Reiman EM, Fievet N, Rotter JI, Reisch JS, Hanon O, Cupidi C, Andre Uitterlinden AG, Royall DR, Dufouil C, Maletta RG, de Rojas I, Sano M, Brice A, Cecchetti R, George-Hyslop PS, Ritchie K, Tsolaki M, Tsuang DW, Dubois B, Craig D, Wu CK, Soininen H, Avramidou D, Albin RL, Fratiglioni L, Germanou A, Apostolova LG, Keller L, Koutroumani M, Arnold SE, Panza F, Gkatzima O, Asthana S, Hannequin D, Whitehead P, Atwood CS, Caffarra P, Hampel H, Quintela I, Carracedo A, Lannfelt L, Rubinsztein DC, Barnes LL, Pasquier F, Frolich L, Barral S, McGuinness B, Beach TG, Johnston JA, Becker JT, Passmore P, Bigio EH, Schott JM, Bird TD, Warren JD, Boeve BF, Lupton MK, Bowen JD, Proitsi P, Boxer A, Powell JF, Burke JR, Kauwe JSK, Burns JM, Mancuso M, Buxbaum JD, Bonuccelli U, Cairns NJ, McQuillin A, Cao C, Livingston G, Carlson CS, Bass NJ, Carlsson CM, Hardy J, Carney RM, Bras J, Carrasquillo MM, Guerreiro R, Allen M, Chui HC, Fisher E, Masullo C, Crocco EA, DeCarli C, Bisceglio G, Dick M, Ma L, Duara R, Graff-Radford NR, Evans DA, Hodges A, Faber KM, Scherer M, Fallon KB, Riemenschneider M, Fardo DW, Heun R, Farlow MR, Kolsch H, Ferris S, Leber M, Foroud TM, Heuser I, Galasko DR, Giegling I, Gearing M, Hull M, Geschwind DH, Gilbert JR, Morris J, Green RC, Mayo K, Growdon JH, Feulner T, Hamilton RL, Harrell LE, Drichel D, Honig LS, Cushion TD, Huentelman MJ, Hollingworth P, Hulette CM, Hyman BT, Marshall R, Jarvik GP, Meggy A, Abner E, Menzies GE, Jin LW, Leonenko G, Real LM, Jun GR, Baldwin CT, Grozeva D, Karydas A, Russo G, Kaye JA, Kim R, Jessen F, Kowall NW, Vellas B, Kramer JH, Vardy E, LaFerla FM, Jockel KH, Lah JJ, Dichgans M, Leverenz JB, Mann D, Levey AI, Pickering-Brown S, Lieberman AP, Klopp N, Lunetta KL, Wichmann HE, Lyketsos CG, Morgan K, Marson DC, Brown K, Martiniuk F, Medway C, Mash DC, Nothen MM, Masliah E, Hooper NM, McCormick WC, Daniele A, McCurry SM, Bayer A, McDavid AN, Gallacher J, McKee AC, van den Bussche H, Mesulam M, Brayne C, Miller BL, Riedel-Heller S, Miller CA, Miller JW, Al-Chalabi A, Morris JC, Shaw CE, Myers AJ, Wiltfang J, O’Bryant S, Olichney JM, Alvarez V, Parisi JE, Singleton AB, Paulson HL, Collinge J, Perry WR, Mead S, Peskind E, Cribbs DH, Rossor M, Pierce A, Ryan NS, Poon WW, Nacmias B, Potter H, Sorbi S, Quinn JF, Sacchinelli E, Raj A, Spalletta G, Raskind M, Caltagirone C, Bossu P, Orfei MD, Reisberg B, Clarke R, Reitz C, Smith AD, Ringman JM, Warden D, Roberson ED, Wilcock G, Rogaeva E, Bruni AC, Rosen HJ, Gallo M, Rosenberg RN, Ben-Shlomo Y, Sager MA, Mecocci P, Saykin AJ, Pastor P, Cuccaro ML, Vance JM, Schneider JA, Schneider LS, Slifer S, Seeley WW, Smith AG, Sonnen JA, Spina S, Stern RA, Swerdlow RH, Tang M, Tanzi RE, Trojanowski JQ, Troncoso JC, Van Deerlin VM, Van Eldik LJ, Vinters HV, Vonsattel JP, Weintraub S, Welsh-Bohmer KA, Wilhelmsen KC, Williamson J, Wingo TS, Woltjer RL, Wright CB, Yu CE, Yu L, Saba Y, Pilotto A, Bullido MJ, Peters O, Crane PK, Bennett D, Bosco P, Coto E, Boccardi V, De Jager PL, Lleo A, Warner N, Lopez OL, Ingelsson M, Deloukas P, Cruchaga C, Graff C, Gwilliam R, Fornage M, Goate AM, Sanchez-Juan P, Kehoe PG, Amin N, Ertekin-Taner N, Berr C, Debette S, Love S, Launer LJ, Younkin SG, Dartigues JF, Corcoran C, Ikram MA, Dickson DW, Nicolas G, Campion D, Tschanz J, Schmidt H, Hakonarson H, Clarimon J, Munger R, Schmidt R, Farrer LA, Van Broeckhoven C, M COD, DeStefano AL, Jones L, Haines JL, Deleuze JF, Owen MJ, Gudnason V, Mayeux R, Escott-Price V, Psaty BM, Ramirez A, Wang LS, Ruiz A, van Duijn CM, Holmans PA, Seshadri S, Williams J, Amouyel P, Schellenberg GD, Lambert JC, Pericak-Vance MA, Alzheimer Disease Genetics C, European Alzheimer’s Disease I, Cohorts for H, Aging Research in Genomic Epidemiology C, Genetic, Environmental Risk in Ad/Defining Genetic P, Environmental Risk for Alzheimer’s Disease C (2019) Genetic meta-analysis of diagnosed Alzheimer’s disease identifies new risk loci and implicates Abeta, tau, immunity and lipid processing. Nat Genet 51, 414–430.

[13] Lambert JC, Ibrahim-Verbaas CA, Harold D, Naj AC, Sims R, Bellenguez C, DeStafano AL, Bis JC, Beecham GW, Grenier-Boley B, Russo G, Thorton-Wells TA, Jones N, Smith AV, Chouraki V, Thomas C, Ikram MA, Zelenika D, Vardarajan BN, Kamatani Y, Lin CF, Gerrish A, Schmidt H, Kunkle B, Dunstan ML, Ruiz A, Bihoreau MT, Choi SH, Reitz C, Pasquier F, Cruchaga C, Craig D, Amin N, Berr C, Lopez OL, De Jager PL, Deramecourt V, Johnston JA, Evans D, Lovestone S, Letenneur L, Moron FJ, Rubinsztein DC, Eiriksdottir G, Sleegers K, Goate AM, Fievet N, Huentelman MW, Gill M, Brown K, Kamboh MI, Keller L, Barberger-Gateau P, McGuiness B, Larson EB, Green R, Myers AJ, Dufouil C, Todd S, Wallon D, Love S, Rogaeva E, Gallacher J, St George-Hyslop P, Clarimon J, Lleo A, Bayer A, Tsuang DW, Yu L, Tsolaki M, Bossu P, Spalletta G, Proitsi P, Collinge J, Sorbi S, Sanchez-Garcia F, Fox NC, Hardy J, Deniz Naranjo MC, Bosco P, Clarke R, Brayne C, Galimberti D, Mancuso M, Matthews F, European Alzheimer’s Disease I, Genetic Environmental Risk in Alzheimer’s D, Alzheimer’s Disease Genetic C, Cohorts for H, Aging Research in Genomic E, Moebus S, Mecocci P, Del Zompo M, Maier W, Hampel H, Pilotto A, Bullido M, Panza F, Caffarra P, Nacmias B, Gilbert JR, Mayhaus M, Lannefelt L, Hakonarson H, Pichler S, Carrasquillo MM, Ingelsson M, Beekly D, Alvarez V, Zou F, Valladares O, Younkin SG, Coto E, Hamilton-Nelson KL, Gu W, Razquin C, Pastor P, Mateo I, Owen MJ, Faber KM, Jonsson PV, Combarros O, O’Donovan MC, Cantwell LB, Soininen H, Blacker D, Mead S, Mosley TH, Jr., Bennett DA, Harris TB, Fratiglioni L, Holmes C, de Bruijn RF, Passmore P, Montine TJ, Bettens K, Rotter JI, Brice A, Morgan K, Foroud TM, Kukull WA, Hannequin D, Powell JF, Nalls MA, Ritchie K, Lunetta KL, Kauwe JS, Boerwinkle E, Riemenschneider M, Boada M, Hiltuenen M, Martin ER, Schmidt R, Rujescu D, Wang LS, Dartigues JF, Mayeux R, Tzourio C, Hofman A, Nothen MM, Graff C, Psaty BM, Jones L, Haines JL, Holmans PA, Lathrop M, Pericak-Vance MA, Launer LJ, Farrer LA, van Duijn CM, Van Broeckhoven C, Moskvina V, Seshadri S, Williams J, Schellenberg GD, Amouyel P (2013) Meta-analysis of 74,046 individuals identifies 11 new susceptibility loci for Alzheimer’s disease. Nat Genet 45, 1452–1458.

[14] Jansen IE, Savage JE, Watanabe K, Bryois J, Williams DM, Steinberg S, Sealock J, Karlsson IK, Hagg S, Athanasiu L, Voyle N, Proitsi P, Witoelar A, Stringer S, Aarsland D, Almdahl IS, Andersen F, Bergh S, Bettella F, Bjornsson S, Braekhus A, Brathen G, de Leeuw C, Desikan RS, Djurovic S, Dumitrescu L, Fladby T, Hohman TJ, Jonsson PV, Kiddle SJ, Rongve A, Saltvedt I, Sando SB, Selbaek G, Shoai M, Skene NG, Snaedal J, Stordal E, Ulstein ID, Wang Y, White LR, Hardy J, Hjerling-Leffler J, Sullivan PF, van der Flier WM, Dobson R, Davis LK, Stefansson H, Stefansson K, Pedersen NL, Ripke S, Andreassen OA, Posthuma D (2019) Genome-wide meta-analysis identifies new loci and functional pathways influencing Alzheimer’s disease risk. Nat Genet 51, 404–413.

[15] Bellenguez C, Kucukali F, Jansen IE, Kleineidam L, Moreno-Grau S, Amin N, Naj AC, Campos-Martin R, Grenier-Boley B, Andrade V, Holmans PA, Boland A, Damotte V, van der Lee SJ, Costa MR, Kuulasmaa T, Yang Q, de Rojas I, Bis JC, Yaqub A, Prokic I, Chapuis J, Ahmad S, Giedraitis V, Aarsland D, Garcia-Gonzalez P, Abdelnour C, Alarcon-Martin E, Alcolea D, Alegret M, Alvarez I, Alvarez V, Armstrong NJ, Tsolaki A, Antunez C, Appollonio I, Arcaro M, Archetti S, Pastor AA, Arosio B, Athanasiu L, Bailly H, Banaj N, Baquero M, Barral S, Beiser A, Pastor AB, Below JE, Benchek P, Benussi L, Berr C, Besse C, Bessi V, Binetti G, Bizarro A, Blesa R, Boada M, Boerwinkle E, Borroni B, Boschi S, Bossu P, Brathen G, Bressler J, Bresner C, Brodaty H, Brookes KJ, Brusco LI, Buiza-Rueda D, Burger K, Burholt V, Bush WS, Calero M, Cantwell LB, Chene G, Chung J, Cuccaro ML, Carracedo A, Cecchetti R, Cervera-Carles L, Charbonnier C, Chen HH, Chillotti C, Ciccone S, Claassen J, Clark C, Conti E, Corma-Gomez A, Costantini E, Custodero C, Daian D, Dalmasso MC, Daniele A, Dardiotis E, Dartigues JF, de Deyn PP, de Paiva Lopes K, de Witte LD, Debette S, Deckert J, Del Ser T, Denning N, DeStefano A, Dichgans M, Diehl-Schmid J, Diez-Fairen M, Rossi PD, Djurovic S, Duron E, Duzel E, Dufouil C, Eiriksdottir G, Engelborghs S, Escott-Price V, Espinosa A, Ewers M, Faber KM, Fabrizio T, Nielsen SF, Fardo DW, Farotti L, Fenoglio C, Fernandez-Fuertes M, Ferrari R, Ferreira CB, Ferri E, Fin B, Fischer P, Fladby T, Fliessbach K, Fongang B, Fornage M, Fortea J, Foroud TM, Fostinelli S, Fox NC, Franco-Macias E, Bullido MJ, Frank-Garcia A, Froelich L, Fulton-Howard B, Galimberti D, Garcia-Alberca JM, Garcia-Gonzalez P, Garcia-Madrona S, Garcia-Ribas G, Ghidoni R, Giegling I, Giorgio G, Goate AM, Goldhardt O, Gomez-Fonseca D, Gonzalez-Perez A, Graff C, Grande G, Green E, Grimmer T, Grunblatt E, Grunin M, Gudnason V, Guetta-Baranes T, Haapasalo A, Hadjigeorgiou G, Haines JL, Hamilton-Nelson KL, Hampel H, Hanon O, Hardy J, Hartmann AM, Hausner L, Harwood J, Heilmann-Heimbach S, Helisalmi S, Heneka MT, Hernandez I, Herrmann MJ, Hoffmann P, Holmes C, Holstege H, Vilas RH, Hulsman M, Humphrey J, Biessels GJ, Jian X, Johansson C, Jun GR, Kastumata Y, Kauwe J, Kehoe PG, Kilander L, Stahlbom AK, Kivipelto M, Koivisto A, Kornhuber J, Kosmidis MH, Kukull WA, Kuksa PP, Kunkle BW, Kuzma AB, Lage C, Laukka EJ, Launer L, Lauria A, Lee CY, Lehtisalo J, Lerch O, Lleo A, Longstreth W, Jr., Lopez O, de Munain AL, Love S, Lowemark M, Luckcuck L, Lunetta KL, Ma Y, Macias J, MacLeod CA, Maier W, Mangialasche F, Spallazzi M, Marquie M, Marshall R, Martin ER, Montes AM, Rodriguez CM, Masullo C, Mayeux R, Mead S, Mecocci P, Medina M, Meggy A, Mehrabian S, Mendoza S, Menendez-Gonzalez M, Mir P, Moebus S, Mol M, Molina-Porcel L, Montrreal L, Morelli L, Moreno F, Morgan K, Mosley T, Nothen MM, Muchnik C, Mukherjee S, Nacmias B, Ngandu T, Nicolas G, Nordestgaard BG, Olaso R, Orellana A, Orsini M, Ortega G, Padovani A, Paolo C, Papenberg G, Parnetti L, Pasquier F, Pastor P, Peloso G, Perez-Cordon A, Perez-Tur J, Pericard P, Peters O, Pijnenburg YAL, Pineda JA, Pinol-Ripoll G, Pisanu C, Polak T, Popp J, Posthuma D, Priller J, Puerta R, Quenez O, Quintela I, Thomassen JQ, Rabano A, Rainero I, Rajabli F, Ramakers I, Real LM, Reinders MJT, Reitz C, Reyes-Dumeyer D, Ridge P, Riedel-Heller S, Riederer P, Roberto N, Rodriguez-Rodriguez E, Rongve A, Allende IR, Rosende-Roca M, Royo JL, Rubino E, Rujescu D, Saez ME, Sakka P, Saltvedt I, Sanabria A, Sanchez-Arjona MB, Sanchez-Garcia F, Juan PS, Sanchez-Valle R, Sando SB, Sarnowski C, Satizabal CL, Scamosci M, Scarmeas N, Scarpini E, Scheltens P, Scherbaum N, Scherer M, Schmid M, Schneider A, Schott JM, Selbaek G, Seripa D, Serrano M, Sha J, Shadrin AA, Skrobot O, Slifer S, Snijders GJL, Soininen H, Solfrizzi V, Solomon A, Song Y, Sorbi S, Sotolongo-Grau O, Spalletta G, Spottke A, Squassina A, Stordal E, Tartan JP, Tarraga L, Tesi N, Thalamuthu A, Thomas T, Tosto G, Traykov L, Tremolizzo L, Tybjaerg-Hansen A, Uitterlinden A, Ullgren A, Ulstein I, Valero S, Valladares O, Broeckhoven CV, Vance J, Vardarajan BN, van der Lugt A, Dongen JV, van Rooij J, van Swieten J, Vandenberghe R, Verhey F, Vidal JS, Vogelgsang J, Vyhnalek M, Wagner M, Wallon D, Wang LS, Wang R, Weinhold L, Wiltfang J, Windle G, Woods B, Yannakoulia M, Zare H, Zhao Y, Zhang X, Zhu C, Zulaica M, Eadb, Gr@Ace, Degesco, Eadi, Gerad, Demgene, FinnGen, Adgc, Charge, Farrer LA, Psaty BM, Ghanbari M, Raj T, Sachdev P, Mather K, Jessen F, Ikram MA, de Mendonca A, Hort J, Tsolaki M, Pericak-Vance MA, Amouyel P, Williams J, Frikke-Schmidt R, Clarimon J, Deleuze JF, Rossi G, Seshadri S, Andreassen OA, Ingelsson M, Hiltunen M, Sleegers K, Schellenberg GD, van Duijn CM, Sims R, van der Flier WM, Ruiz A, Ramirez A, Lambert JC (2022) New insights into the genetic etiology of Alzheimer’s disease and related dementias. Nat Genet 54, 412–436.

[16] Escott-Price V, Sims R, Bannister C, Harold D, Vronskaya M, Majounie E, Badarinarayan N, Gerad/Perades, consortia I, Morgan K, Passmore P, Holmes C, Powell J, Brayne C, Gill M, Mead S, Goate A, Cruchaga C, Lambert JC, van Duijn C, Maier W, Ramirez A, Holmans P, Jones L, Hardy J, Seshadri S, Schellenberg GD, Amouyel P, Williams J (2015) Common polygenic variation enhances risk prediction for Alzheimer’s disease. Brain 138, 3673–3684.

[17] Jack CR, Jr., Bennett DA, Blennow K, Carrillo MC, Dunn B, Haeberlein SB, Holtzman DM, Jagust W, Jessen F, Karlawish J, Liu E, Molinuevo JL, Montine T, Phelps C, Rankin KP, Rowe CC, Scheltens P, Siemers E, Snyder HM, Sperling R, Contributors (2018) NIA-AA Research Framework: Toward a biological definition of Alzheimer’s disease. Alzheimers Dement 14, 535–562.

[18] Dubois B, Villain N, Frisoni GB, Rabinovici GD, Sabbagh M, Cappa S, Bejanin A, Bombois S, Epelbaum S, Teichmann M, Habert M-O, Nordberg A, Blennow K, Galasko D, Stern Y, Rowe CC, Salloway S, Schneider LS, Cummings JL, Feldman HH (2021) Clinical diagnosis of Alzheimer’s disease: recommendations of the International Working Group. The Lancet Neurology.

[19] Dubois B, Feldman HH, Jacova C, Cummings JL, Dekosky ST, Barberger-Gateau P, Delacourte A, Frisoni G, Fox NC, Galasko D, Gauthier S, Hampel H, Jicha GA, Meguro K, O’Brien J, Pasquier F, Robert P, Rossor M, Salloway S, Sarazin M, de Souza LC, Stern Y, Visser PJ, Scheltens P (2010) Revising the definition of Alzheimer’s disease: a new lexicon. Lancet Neurol 9, 1118–1127.

[20] Fiksinski AM, Hoftman GD, Vorstman JAS, Bearden CE (2023) A genetics-first approach to understanding autism and schizophrenia spectrum disorders: the 22q11.2 deletion syndrome. Mol Psychiatry 28, 341–353.

[21] Stessman HA, Bernier R, Eichler EE (2014) A genotype-first approach to defining the subtypes of a complex disease. Cell 156, 872–877.

[22] Wilczewski CM, Obasohan J, Paschall JE, Zhang S, Singh S, Maxwell GL, Similuk M, Wolfsberg TG, Turner C, Biesecker LG, Katz AE (2023) Genotype first: Clinical genomics research through a reverse phenotyping approach. Am J Hum Genet 110, 3–12.

[23] Neff RA, Wang M, Vatansever S, Guo L, Ming C, Wang Q, Wang E, Horgusluoglu-Moloch E, Song WM, Li A, Castranio EL, Tcw J, Ho L, Goate A, Fossati V, Noggle S, Gandy S, Ehrlich ME, Katsel P, Schadt E, Cai D, Brennand KJ, Haroutunian V, Zhang B (2021) Molecular subtyping of Alzheimer’s disease using RNA sequencing data reveals novel mechanisms and targets. Sci Adv 7, eabb5398.

[24] Udler MS, Kim J, von Grotthuss M, Bonas-Guarch S, Cole JB, Chiou J, Christopher DAoboM, the I, Boehnke M, Laakso M, Atzmon G, Glaser B, Mercader JM, Gaulton K, Flannick J, Getz G, Florez JC (2018) Type 2 diabetes genetic loci informed by multi-trait associations point to disease mechanisms and subtypes: A soft clustering analysis. PLoS Med 15, e1002654.

[25] Arnedo J, Svrakic DM, Del Val C, Romero-Zaliz R, Hernandez-Cuervo H, Molecular Genetics of Schizophrenia C, Fanous AH, Pato MT, Pato CN, de Erausquin GA, Cloninger CR, Zwir I (2015) Uncovering the hidden risk architecture of the schizophrenias: confirmation in three independent genome-wide association studies. Am J Psychiatry 172, 139–153.

[26] Dahl A, Zaitlen N (2020) Genetic Influences on Disease Subtypes. Annu Rev Genomics Hum Genet 21, 413–435.

[27] Rangan AV, McGrouther CC, Kelsoe J, Schork N, Stahl E, Zhu Q, Krishnan A, Yao V, Troyanskaya O, Bilaloglu S, Raghavan P, Bergen S, Jureus A, Landen M, Bipolar Disorders Working Group of the Psychiatric Genomics C (2018) A loop-counting method for covariate-corrected low-rank biclustering of gene-expression and genome-wide association study data. PLoS Comput Biol 14, e1006105.

[28] Sudlow C, Gallacher J, Allen N, Beral V, Burton P, Danesh J, Downey P, Elliott P, Green J, Landray M, Liu B, Matthews P, Ong G, Pell J, Silman A, Young A, Sprosen T, Peakman T, Collins R (2015) UK biobank: an open access resource for identifying the causes of a wide range of complex diseases of middle and old age. PLoS Med 12, e1001779.

[29] Bycroft C, Freeman C, Petkova D, Band G, Elliott LT, Sharp K, Motyer A, Vukcevic D, Delaneau O, O’Connell J, Cortes A, Welsh S, Young A, Effingham M, McVean G, Leslie S, Allen N, Donnelly P, Marchini J (2018) The UK Biobank resource with deep phenotyping and genomic data. Nature 562, 203–209.

[30] Wilkinson T, Schnier C, Bush K, Rannikmae K, Henshall DE, Lerpiniere C, Allen NE, Flaig R, Russ TC, Bathgate D, Pal S, O’Brien JT, Sudlow CLM, Dementias Platform UK, Biobank UK (2019) Identifying dementia outcomes in UK Biobank: a validation study of primary care, hospital admissions and mortality data. Eur J Epidemiol 34, 557–565.

[31] Chen CY, Pollack S, Hunter DJ, Hirschhorn JN, Kraft P, Price AL (2013) Improved ancestry inference using weights from external reference panels. Bioinformatics 29, 1399–1406.

[32] The 1000 Genomes Project Consortium (2015) A global reference for human genetic variation. Nature 526, 68–74.

[33] Das S, Forer L, Schonherr S, Sidore C, Locke AE, Kwong A, Vrieze SI, Chew EY, Levy S, McGue M, Schlessinger D, Stambolian D, Loh PR, Iacono WG, Swaroop A, Scott LJ, Cucca F, Kronenberg F, Boehnke M, Abecasis GR, Fuchsberger C (2016) Next-generation genotype imputation service and methods. Nat Genet 48, 1284–1287.

[34] Chung JE, Magland JF, Barnett AH, Tolosa VM, Tooker AC, Lee KY, Shah KG, Felix SH, Frank LM, Greengard LF (2017) A Fully Automated Approach to Spike Sorting. Neuron 95, 1381–1394 e1386.

[35] Prive F, Luu K, Blum MGB, McGrath JJ, Vilhjalmsson BJ (2020) Efficient toolkit implementing best practices for principal component analysis of population genetic data. Bioinformatics 36, 4449–4457.

[36] (!!! INVALID CITATION !!! (27)).

[37] Zhou H, Lin W, Labra SR, Lipton SA, Schork NJ, Rangan AV (2022) Detecting boolean asymmetric relationships with a loop counting technique and its implications for analyzing heterogeneity within gene expression datasets. bioRxiv, 2022.2008.2004.502792.

[38] Chang CC, Chow CC, Tellier LC, Vattikuti S, Purcell SM, Lee JJ (2015) Second-generation PLINK: rising to the challenge of larger and richer datasets. Gigascience 4, 7.

[39] Raudvere U, Kolberg L, Kuzmin I, Arak T, Adler P, Peterson H, Vilo J (2019) g:Profiler: a web server for functional enrichment analysis and conversions of gene lists (2019 update). Nucleic Acids Res 47, W191–W198.

[40] Wu T, Hu E, Xu S, Chen M, Guo P, Dai Z, Feng T, Zhou L, Tang W, Zhan L, Fu X, Liu S, Bo X, Yu G (2021) clusterProfiler 4.0: A universal enrichment tool for interpreting omics data. Innovation (Camb) 2, 100141.

[41] Yu G, Wang LG, Han Y, He QY (2012) clusterProfiler: an R package for comparing biological themes among gene clusters. OMICS 16, 284–287.

[42] Ashburner M, Ball CA, Blake JA, Botstein D, Butler H, Cherry JM, Davis AP, Dolinski K, Dwight SS, Eppig JT, Harris MA, Hill DP, Issel-Tarver L, Kasarskis A, Lewis S, Matese JC, Richardson JE, Ringwald M, Rubin GM, Sherlock G (2000) Gene ontology: tool for the unification of biology. The Gene Ontology Consortium. Nat Genet 25, 25–29.

[43] Gene Ontology C (2021) The Gene Ontology resource: enriching a GOld mine. Nucleic Acids Res 49, D325–D334.

[44] Merico D, Isserlin R, Stueker O, Emili A, Bader GD (2010) Enrichment map: a network-based method for gene-set enrichment visualization and interpretation. PLoS One 5, e13984.

[45] Febrero M, Galeano P, González-Manteiga W (2007) Outlier detection in functional data by depth measures, with application to identify abnormal NOx levels. Environmetrics 19, 331–345.

[46] Donohue MC, Sperling RA, Petersen R, Sun CK, Weiner MW, Aisen PS, Alzheimer’s Disease Neuroimaging I (2017) Association Between Elevated Brain Amyloid and Subsequent Cognitive Decline Among Cognitively Normal Persons. JAMA 317, 2305–2316.

[47] Donohue MC, Sperling RA, Salmon DP, Rentz DM, Raman R, Thomas RG, Weiner M, Aisen PS, Australian Imaging B, Lifestyle Flagship Study of A, Alzheimer’s Disease Neuroimaging I, Alzheimer’s Disease Cooperative S (2014) The preclinical Alzheimer cognitive composite: measuring amyloid-related decline. JAMA Neurol 71, 961–970.

[48] Landau SM, Breault C, Joshi AD, Pontecorvo M, Mathis CA, Jagust WJ, Mintun MA, Alzheimer’s Disease Neuroimaging I (2013) Amyloid-beta imaging with Pittsburgh compound B and florbetapir: comparing radiotracers and quantification methods. J Nucl Med 54, 70–77.

[49] Landau SM, Marks SM, Mormino EC, Rabinovici GD, Oh H, O’Neil JP, Wilson RS, Jagust WJ (2012) Association of lifetime cognitive engagement and low β-amyloid deposition. Archives of Neurology 69, 623–629.

[50] Shaw LM, Vanderstichele H, Knapik-Czajka M, Clark CM, Aisen PS, Petersen RC, Blennow K, Soares H, Simon A, Lewczuk P, Dean R, Siemers E, Potter W, Lee VM, Trojanowski JQ, Alzheimer’s Disease Neuroimaging I (2009) Cerebrospinal fluid biomarker signature in Alzheimer’s disease neuroimaging initiative subjects. Ann Neurol 65, 403–413.

[51] Bates D, Machler M, Bolker BM, Walker SC (2015) Fitting Linear Mixed-Effects Models Using lme4. Journal of Statistical Software 67, 1–48.

[52] Kuznetsova A, Brockhoff PB, Christensen RHB (2017) lmerTest Package: Tests in Linear Mixed Effects Models. Journal of Statistical Software 82, 1–26.

[53] Stefansson H, Helgason A, Thorleifsson G, Steinthorsdottir V, Masson G, Barnard J, Baker A, Jonasdottir A, Ingason A, Gudnadottir VG, Desnica N, Hicks A, Gylfason A, Gudbjartsson DF, Jonsdottir GM, Sainz J, Agnarsson K, Birgisdottir B, Ghosh S, Olafsdottir A, Cazier JB, Kristjansson K, Frigge ML, Thorgeirsson TE, Gulcher JR, Kong A, Stefansson K (2005) A common inversion under selection in Europeans. Nat Genet 37, 129–137.

54. Pittman AM, Fung HC, de Silva R (2006) Untangling the tau gene association with neurodegenerative disorders. Hum Mol Genet 15 **Spec No 2**, R188–195.

[55] Sims R, Hill M, Williams J (2020) The multiplex model of the genetics of Alzheimer’s disease. Nat Neurosci 23, 311–322.

[56] Price AL, Patterson NJ, Plenge RM, Weinblatt ME, Shadick NA, Reich D (2006) Principal components analysis corrects for stratification in genome-wide association studies. Nat Genet 38, 904–909.

[57] Allen M, Kachadoorian M, Quicksall Z, Zou F, Chai HS, Younkin C, Crook JE, Pankratz VS, Carrasquillo MM, Krishnan S, Nguyen T, Ma L, Malphrus K, Lincoln S, Bisceglio G, Kolbert CP, Jen J, Mukherjee S, Kauwe JK, Crane PK, Haines JL, Mayeux R, Pericak-Vance MA, Farrer LA, Schellenberg GD, Parisi JE, Petersen RC, Graff-Radford NR, Dickson DW, Younkin SG, Ertekin-Taner N (2014) Association of MAPT haplotypes with Alzheimer’s disease risk and MAPT brain gene expression levels. Alzheimers Res Ther 6, 39.

[58] Strickland SL, Reddy JS, Allen M, N’Songo A, Burgess JD, Corda MM, Ballard T, Wang X, Carrasquillo MM, Biernacka JM, Jenkins GD, Mukherjee S, Boehme K, Crane P, Kauwe JS, Ertekin-Taner N, Alzheimer’s Disease Genetics C (2020) MAPT haplotype-stratified GWAS reveals differential association for AD risk variants. Alzheimers Dement 16, 983–1002.

[59] Baker M, Litvan I, Houlden H, Adamson J, Dickson D, Perez-Tur J, Hardy J, Lynch T, Bigio E, Hutton M (1999) Association of an extended haplotype in the tau gene with progressive supranuclear palsy. Hum Mol Genet 8, 711–715.

[60] Munoz L, Ammit AJ (2010) Targeting p38 MAPK pathway for the treatment of Alzheimer’s disease. Neuropharmacology 58, 561–568.

[61] Leyns CEG, Holtzman DM (2017) Glial contributions to neurodegeneration in tauopathies. Mol Neurodegener 12, 50.

[62] Tooyama I, Kimura H, Akiyama H, McGeer PL (1990) Reactive microglia express class I and class II major histocompatibility complex antigens in Alzheimer’s disease. Brain Res 523, 273–280.

[63] Nobes CD, Hall A (1999) Rho GTPases control polarity, protrusion, and adhesion during cell movement. J Cell Biol 144, 1235–1244.

64. Settleman J (1999) Rho GTPases in Development In Cytoskeleton and Small G Proteins, Jeanteur P, ed. Springer Berlin Heidelberg, Berlin, Heidelberg, pp. 201–229.

[65] Hotulainen P, Hoogenraad CC (2010) Actin in dendritic spines: connecting dynamics to function. J Cell Biol 189, 619–629.

[66] Cabrales Fontela Y, Kadavath H, Biernat J, Riedel D, Mandelkow E, Zweckstetter M (2017) Multivalent cross-linking of actin filaments and microtubules through the microtubule-associated protein Tau. Nat Commun 8, 1981.

[67] Valenta T, Hausmann G, Basler K (2012) The many faces and functions of beta-catenin. EMBO J 31, 2714–2736.

[68] Soldano A, Hassan BA (2014) Beyond pathology: APP, brain development and Alzheimer’s disease. Curr Opin Neurobiol 27, 61–67.

[69] Kowall NW, Kosik KS (1987) Axonal disruption and aberrant localization of tau protein characterize the neuropil pathology of Alzheimer’s disease. Ann Neurol 22, 639–643.

[70] Iqbal K, Liu F, Gong CX, Grundke-Iqbal I (2010) Tau in Alzheimer disease and related tauopathies. Curr Alzheimer Res 7, 656–664.

71. Schork NJ, Elman JA (2023) Pathway-specific polygenic risk scores correlate with clinical status and Alzheimer’s-related biomarkers. Res Sq.

72. La Joie R, Visani AV, Baker SL, Brown JA, Bourakova V, Cha J, Chaudhary K, Edwards L, Iaccarino L, Janabi M, Lesman-Segev OH, Miller ZA, Perry DC, O’Neil JP, Pham J, Rojas JC, Rosen HJ, Seeley WW, Tsai RM, Miller BL, Jagust WJ, Rabinovici GD (2020) Prospective longitudinal atrophy in Alzheimer’s disease correlates with the intensity and topography of baseline tau-PET. Sci Transl Med 12, eaau5732.

[73] Hanseeuw BJ, Betensky RA, Jacobs HIL, Schultz AP, Sepulcre J, Becker JA, Cosio DMO, Farrell M, Quiroz YT, Mormino EC, Buckley RF, Papp KV, Amariglio RA, Dewachter I, Ivanoiu A, Huijbers W, Hedden T, Marshall GA, Chhatwal JP, Rentz DM, Sperling RA, Johnson K (2019) Association of Amyloid and Tau With Cognition in Preclinical Alzheimer Disease: A Longitudinal Study. JAMA Neurol 76, 915–924.

74. Ossenkoppele R, Lyoo CH, Sudre CH, van Westen D, Cho H, Ryu YH, Choi JY, Smith R, Strandberg O, Palmqvist S, Westman E, Tsai R, Kramer J, Boxer AL, Gorno-Tempini ML, La Joie R, Miller BL, Rabinovici GD, Hansson O (2020) Distinct tau PET patterns in atrophy-defined subtypes of Alzheimer’s disease. Alzheimers Dement 16, 335–344.

[75] Saporta AS, Sottile SL, Miller LJ, Feely SM, Siskind CE, Shy ME (2011) Charcot-Marie-Tooth disease subtypes and genetic testing strategies. Ann Neurol 69, 22–33.

[76] Crutch SJ, Schott JM, Rabinovici GD, Murray M, Snowden JS, van der Flier WM, Dickerson BC, Vandenberghe R, Ahmed S, Bak TH, Boeve BF, Butler C, Cappa SF, Ceccaldi M, de Souza LC, Dubois B, Felician O, Galasko D, Graff-Radford J, Graff-Radford NR, Hof PR, Krolak-Salmon P, Lehmann M, Magnin E, Mendez MF, Nestor PJ, Onyike CU, Pelak VS, Pijnenburg Y, Primativo S, Rossor MN, Ryan NS, Scheltens P, Shakespeare TJ, Suarez Gonzalez A, Tang-Wai DF, Yong KXX, Carrillo M, Fox NC, Alzheimer’s Association IAAsD, Associated Syndromes Professional Interest A (2017) Consensus classification of posterior cortical atrophy. Alzheimers Dement 13, 870–884.

[77] Gorno-Tempini ML, Hillis AE, Weintraub S, Kertesz A, Mendez M, Cappa SF, Ogar JM, Rohrer JD, Black S, Boeve BF, Manes F, Dronkers NF, Vandenberghe R, Rascovsky K, Patterson K, Miller BL, Knopman DS, Hodges JR, Mesulam MM, Grossman M (2011) Classification of primary progressive aphasia and its variants. Neurology 76, 1006–1014.

[78] Townley RA, Graff-Radford J, Mantyh WG, Botha H, Polsinelli AJ, Przybelski SA, Machulda MM, Makhlouf AT, Senjem ML, Murray ME, Reichard RR, Savica R, Boeve BF, Drubach DA, Josephs KA, Knopman DS, Lowe VJ, Jack CR, Jr., Petersen RC, Jones DT (2020) Progressive dysexecutive syndrome due to Alzheimer’s disease: a description of 55 cases and comparison to other phenotypes. Brain Commun 2, fcaa068.

[79] Ossenkoppele R, Singleton EH, Groot C, Dijkstra AA, Eikelboom WS, Seeley WW, Miller B, Laforce RJ, Scheltens P, Papma JM, Rabinovici GD, Pijnenburg YAL (2022) Research Criteria for the Behavioral Variant of Alzheimer Disease: A Systematic Review and Meta-analysis. JAMA Neurol 79, 48–60.

[80] (!!! INVALID CITATION !!! (12)).

