## Supplementary Material for "Exploring the genetic heterogeneity of Alzheimer’s disease: Evidence for genetic subtypes"

### Supplementary Methods

#### *Rationale for allele-coding*

Our biclustering-analysis is applied after encoding the genetic-data using allele-combinations. Specifically, each SNP is associated with three values: homozygous-dominant (AA), heterozygous (Aa) and homozygous-recessive (aa). These possible values are encoded within three SNP-specific columns of our data-matrix. Thus, a particular participant who is homozygous-dominant for a particular SNP will have corresponding column-values of [1, 0, 0], a participant who is heterozygous for that SNP would have corresponding column-values of [0, 1, 0], and a participant who is homozygous-recessive for that SNP would have column-values of [0, 0, 1]. This 'allele-coding' is an alternative to the more commonly used 'additive-coding', which records the number of rare-variants per SNP (i.e., 0, 1 or 2, also known as 'dosage-coding'). Allele-coded data contains the same information as additively-coded data: no information is lost, and one can easily transform one encoding-paradigm into the other. Nevertheless, allele-coding can be advantageous when searching for certain kinds of structure within genetic-data. As a first simple example, imagine a single disease-relevant SNP with the following (idealistic) distribution:

| | AA | Aa | aa |
|---|---|---|---|
| population-1: | 24% | 52% | 24% |
| population-2: | 26% | 48% | 26% |

When additively-coded, this SNP has the same mean but different variances across the two populations. Assuming that population-1 corresponds to controls and population-2 to cases, identifying such an additively-coded SNP as disease-relevant would require looking for 'second-order' effects (i.e., higher-order moments of the data such as the variance). By contrast, when allele-coded, this SNP can be identified as disease-relevant using only first-order statistics (such as population-averages). The above example only considers a single SNP and assumes homogeneity across the population. In the presence of multiple SNPs and/or heterogeneity, the differences between additive- and allele-coding can be even more stark.

As a second example, consider two SNPs, A and B, distributed as follows:

| pop0 | AA | Aa | aa |
|---|---|---|---|
| BB | 2.48% | 9.21% | 8.56% |
| Bb | 6.06% | 22.52% | 20.91% |
| bb | 3.71% | 13.76% | 12.78% |

| pop1 | AA | Aa | aa |
|---|---|---|---|
| BB | 2.35% | 5.61% | 12.29% |
| Bb | 9.90% | 22.59% | 17.01% |
| bb | 0.00% | 17.30% | 12.95% |

| pop2 | AA | Aa | aa |
|---|---|---|---|
| BB | 1.03% | 14.02% | 5.20% |
| Bb | 11.22% | 8.40% | 29.88% |
| bb | 0.00% | 23.08% | 7.17% |

| pop3 | AA | Aa | aa |
|---|---|---|---|
| BB | 0.00% | 12.00% | 8.25% |
| Bb | 8.68% | 21.63% | 19.19% |
| bb | 3.57% | 11.87% | 14.82% |

In each of these tables the rows correspond to the allele-combinations AA, Aa and aa, while the columns correspond to the allele-combinations BB, Bb and bb. Each cell indicates the joint-probability of observing a participant with the corresponding pair of allele-combinations. These joint-distributions illustrate one idealized example of the following phenomenon. For each population:

1. the percentages add up to 100%
2. SNP-A has an (arbitrary) fixed minor-allele-frequency (maf) of $0.35 = 35\%$
3. SNP-A has an additively-coded variance of $2 \cdot maf_A \cdot (1 - maf_A) = 0.455$
4. SNP-B also has a fixed arbitrary maf, in this scenario $0.45$.

5. SNP-B has an additively-coded variance of $0.495$.

6. the correlation between SNP-A and SNP-B is 0 (under additive-coding).

Thus, the 9 numbers associated with each of the joint-distributions above are subject to 6 constraints, yielding a (9-6)=3 dimensional landscape of `freedom' in which the joint-distributions can vary without affecting these constraints. Within this landscape one might imagine disease-specific heterogeneity: Population-0 might represent the control-population (i.e., participants that do not exhibit the disease), while the remaining populations could describe the cases (i.e., those exhibiting the disease). In this scenario, populations 1, 2 and 3 correspond to different subsets of the case-population exhibiting different genetic structure (i.e., disease-specific LD between snps A and B). One goal of a biclustering algorithm would be to expose this heterogeneity by identifying populations 1, 2 and 3 from within the overall case-population. With this goal in mind, we see a clear difference between additive- and allele-coding. The correlation between the additively-coded columns for population-1 (i.e., one of the case-biclusters) is essentially 0, just like the correlation between the same columns for populations 2 and 3 (i.e., the other case-biclusters). Meanwhile, if we use allele-coding, the correlation between the aa- and Bb-columns for population-1 is strongly negative (about -0.17), while the correlation between the same columns in population-2 is strongly positive (about 0.37) and the analogous correlation for population-3 is closer to zero (about -0.06). Several other column-pairs exhibit a similar phenomenon, and the collection of allele-coded correlations differs substantially across the populations.

In sum, for this example, a correlation-based biclustering algorithm won't typically be able to distinguish these populations within the additively-coded data, but will easily be able to distinguish the populations (and correctly identify the heterogeneity) within the allele-coded data. This example is rather idealized, with the joint-probabilities specifically chosen so that they are quite far away from the control-distribution (while remaining on the admissible landscape described above). Nevertheless, one can imagine less extreme scenarios exhibiting similar phenomena: there are many kinds of disease-specific heterogeneity (involving subtle correlations between allele-combinations) which could be more easily accessed via allele-coding than by additive-coding.

Reasoning along the lines above has informed our methodology. In general, there are many kinds of data-encoding one might consider, ranging from simple to very complicated, and different encoding-paradigms may render certain kinds of information more easily accessible to certain kinds of analysis. Ultimately, we are interested in searching for biclusters defined in terms of shared genetic structure. Due to computational-limitations, the analytical tools we have access to are essentially limited to measuring conditional averages and correlations (i.e., estimating low-order moments of the unknown joint-distributions). Because of these limitations, we prefer allele-coding to additive-coding, as the former allows certain kinds of biclusters to be detected using the low-order moments we can access.

***Searching for disease-specific biclusters***

To search for biclusters we use the half-loop method described in Rangan et al. [1]. This strategy involves an iterative process which starts with all the participants (in this case, all participants in a given constellation) and SNPs, and then sequentially removes AD cases and allele-combinations from consideration. Briefly, we can take an $M_D \times N$ matrix of cases $D$ and an $M_X \times N$ matrix of controls $X$. The entry $D_{j,k}$ records the presence or absence of the $k$-th allele-combination for the $j^{\text{th}}$ case, and the entry $X_{j,k}$ records the presence or absence of the $k^{\text{th}}$ allele-combination for the $j^{\text{th}}$ control. For each case $j$ we can measure the fraction of cases (i.e., restricted to matrix $D$) that share the allele-combination $k$ and call this $[D \leftrightarrow D]_{j,k}$. Similarly, we can calculate the fraction of controls that share the allele-combination $k$ and call this $[D \leftrightarrow X]_{j,k}$ (i.e., extending into matrix $X$). We can then calculate $Z_{j,k} = [D \leftrightarrow D]_{j,k} - [D \leftrightarrow X]_{j,k}$, which represents the difference in fraction of shared allele-combinations between the $j^{\text{th}}$ case and other remaining cases compared to the fraction shared with controls for the $k^{\text{th}}$ allele-combination. Averaging over the remaining allele-combinations will give us $Z_j$, referred to as row-scores in our algorithm. Finally, averaging over the remaining cases will leave us with $Z$, or the row trace, which indicates the disease-related signal strength for the remaining subset of AD cases and allele-combinations. At each iteration the cases and allele-combinations with the smallest contributions to $Z$ are removed and the row trace is recalculated as described above.

This process is conducted while controlling for the first two genetic principal-components. Covariate correction is also described in detail in Rangan et al. [1], but essentially weights the contributions of cases and allele-combinations such that the row score and row trace (as well as analogous column-scores and column trace) will be shrunk if drawn from an imbalanced distribution of the covariate space. That is, the biclustering algorithm will tend to ignore these structures in favor of structures that are more evenly distributed in covariate space.

As a null-hypothesis, we assume that the disease-label (i.e., case vs control) is not associated with the genetic profile of each participant. We can draw samples from this null-hypothesis by randomly permuting the case- and control-labels across participants with similar genome-wide principal components (see Rangan et al. [1] for details). Here, we use 500 permutations. By comparing the original trace with the distribution of traces drawn from the null-hypothesis, we can assign a p-value to the observed trace $Z$ at each iteration. In this case, we only assess iterations that include >5% of the cases (i.e., the final iterations with very few cases and/or variants are ignored). To determine whether we have found a statistically significant bicluster within the original data (termed the 'dominant' bicluster), we can examine the p-value of the highest peak ($p_{max}$) and the average p-value across iterations ($p_{avg}$). Depending on the structure of the bicluster, the various p-values may be quite different, but each may be a useful metric for identifying significant biclusters. For example, if the original trace has one or more clear peaks (as shown in **Fig. 3A**), then there are statistically robust 'cutpoints' which can be used to delineate bicluster membership and $p_{max}$ is likely to be very small. On the other hand, if the original trace has a very broad peak or a long plateau (as shown in **Fig. 3B**), then the bicluster is quite 'fuzzy', corresponding to a continuum of membership. In this case, we might expect $p_{avg}$ to be small, but $p_{max}$ may be relatively large. Moreover, we can use the peak of the original trace to delineate the membership of the dominant bicluster (i.e., which AD cases and allellic-combinations contribute to the disease-specific signal). We identify the peak by finding the internal maximum, ignoring the initial and final iterations that include >95% or <5% of AD cases.

If the dominant bicluster within a data-set is statistically significant, we can extract it and then search for a secondary bicluster. This is done by scrambling the entries of the submatrix associated with the bicluster (i.e., entries corresponding to the participants and allele-combinations that were retained in the bicluster) and running the search algorithm again [1, 2].

#### *Replication of disease-relevant constellations in ADNI*

We assessed the extent to which the two levels of structure found in the UKB data were also present in the ADNI data. The first level of the hierarchical structure observed in the UKB data involved the constellations shown in **Fig. 1**. To assess the replication of these structures within the ADNI data, we determined the subset of SNPs which lie in the intersection of the UKB and ADNI datasets, and then further restricted this subset of SNPs to include only those SNPs which had Kunkle p-values of <0.05, denoting this restricted subset by $S_\cap$. Mirroring the original analysis, we then re-calculated the dominant 2 principal-components for the UKB data across $S_\cap$ to obtain loadings $u_1^{UKB}$ and $u_2^{UKB}$ or each participant, and loadings $v1$ and $v2$ for each SNP. Next, we use $v1$ and $v2$ to calculate projections $u_1^{ADNI}$ and $u_2^{ADNI}$ for each ADNI participant. We can then compare the distribution of $u_1^{UKB}$ and $u_2^{UKB}$ with the distribution of $u_1^{ADNI}$ and $u_2^{ADNI}$ to assess overlap between datasets. ADNI participants were assigned to the nearest constellation based on participant loadings on the first principal component.

***Replication of disease-specific biclusters in ADNI***

After participants were classified into constellations, we assessed replication of the dominant bicluster separately in the two constellations in which they were found. First, we restrict the subset of SNPs delineating the bicluster to those that overlap with the ADNI data, referring to that (restricted) subset as $\mathcal{K}$, which is itself a subset of $S_\cap$. We can organize the data into a $|\mathcal{J}| \times |\mathcal{K}|$ array $A^{UKB}$, where $\mathcal{J}$ represents only UKB participants from a given constellation. Likewise, the ADNI data can be organized into a $|\mathcal{J}'| \times |\mathcal{K}|$ array where $\mathcal{J}'$ represents the ADNI participants that have been assigned to the constellation of interest. The bicluster itself, denoted by $B$, is a $|\mathcal{J}_B| \times |\mathcal{K}|$ submatrix of $A^{UKB}$. We then calculate the dominant SNP-wise principal components $v_1$ and $v_2$ of $B$ across $\mathcal{K}$. We use SNP loadings $v_1$ and $v_2$ to calculate projections for each of the $\mathcal{J}$ UKB participants in the constellation and defined $u_1^{UKB}$ and $u_2^{UKB}$ as the first two columns (i.e., participant-wise principal components). Similarly, we can calculate $u_1^{ADNI}$ and $u_2^{ADNI}$ for each of the $\mathcal{J}'$ ADNI participants belonging to the constellation.

After projecting UKB and ADNI participants onto the dominant principal components of $\mathcal{K}$ we must first align the distributions of $u_1^{ADNI}$ and $u_2^{ADNI}$ with $u_1^{UKB}$ and $u_2^{UKB}$. To do so, we use a point-set registration algorithm described in the section "*Affine point-set registration*" to calculate an affine transform that can be applied to perform this alignment.

Following alignment of the two distributions, we use the methods described in the section "*Calculating label similarity*" to calculate case-control label similarity of participants in the ADNI dataset to nearby participants in the UKB dataset, $\tilde{\sigma}(f;\{y_j\},\{l_j\},\{y'_{j'}\},\{l'_{j'}\})$. In this expression, $y_j$ and $l_j$ refer to the projected data $(u_1^{UKB}, u_2^{UKB})$ and case-control label for UKB participant $j$, respectively. The terms $y'_{j'}$ and $l'_{j'}$ refer to the projected and aligned data $\left(u_1^{ADNI}, u_2^{ADNI}\right)$ and case-control label for ADNI participant $j'$, respectively. The value $f$ represents the fraction of individuals from the training set (i.e., UKB data) one should use as nearest neighbors when averaging to approximate the true label of a point in the testing set (e.g., ADNI data).

We can assess the statistical significance of the similarity measure $\tilde{\sigma}\left(f;\{y_j\},\{l_j\},\left\{y'_{j'}\right\},\{l'_{j'}\}\right)$ with a permutation test. The null-hypothesis is that the labels are independent from the position of each point in projected space. Thus, we can draw a sample from the null-hypothesis by (i) randomly permuting the labels $l_j$ within the UKB data, (ii) randomly permuting the labels $l'_{j'}$ within the ADNI data, and then (iii) recalculating the above steps for this label-shuffled data, including the affine point-set registration. Here, we ran 500 permutations. After accumulating a distribution of values for $\tilde{\sigma}(f)$ under the null-hypothesis, we can compare the original $\tilde{\sigma}(f)$ to this distribution. To estimate this empirical p-value, we first calculate the average $z$-score across a range of $f$ in $(f_{lo}, f_{hi})$. For $f_{lo}$ we choose the lower end of the 95% confidence-interval for affine-point-matching, determined using different random initializations applied to random projections of the data, and for $f_{hi}$ we choose 50%. This average $z$-score is calculated using a normalizing factor to correct for heteroskedasticity [3]. The variance determining the normalizing factor is calculated from the analogous $z$-scores obtained after alignment of the projections onto principal-components calculated from randomly selected biclusters (i.e., subsets of cases and allele-combinations) of the same size as the bicluster of interest. We can calculate a global empirical p-value by comparing the average $z$-score of the observed data across the range of parameter choices $f$ in $(f_{lo}, f_{hi})$ to the null distribution across the same range. The empirical p-values reported in the main-text are robust to changes in $f_{lo}$ and $f_{hi}$ (e.g., we get essentially the same p-values if $f_{lo}$ is varied within the range $[1\%, 10\%]$). Given that a particular bicluster is globally significant, values of $f$ corresponding to high $z$-scores (or ranking relative to the null distribution) indicate reasonable values of $f$ to use when labelling new data. This use-case is further described in the main-text.

***Affine point-set registration***

Prior to calculating label similarity between datasets, we perform affine point set registration. In the scenario where the dimension of the data (i.e., number of SNPs) is much larger than the number of samples, the principal components may be overfit to the training dataset. That is, the principal components are calculated to maximize the variance in the training set but not the test set. When projecting the test set data onto the principal components of the training set, the participant data from the test set will almost certainly have a smaller amplitude than the participant data in the training set. Technical artifacts, such as might be introduced by different genotyping chips across studies, may also lead to shifts or distortions in the data distributions. Given two collections of points, say in $\mathbb{R}^2$, we would therefore like to construct a transformation $T : \mathbb{R}^2 \rightarrow \mathbb{R}^2$ which attempts to align one set of points to the other. The strategy we employ (described below) is designed to be part of our bicluster replication pipeline. Thus, we restrict ourselves to affine linear transformations, with the goal of registering the projected values of one dataset to the projected values of another, while controlling for possible differences in the number of cases and controls between these two datasets. While our algorithm may not be 'optimal', it has certain advantages. Specifically, our algorithm is 'automatic', and there are no free parameters which need to be tuned.

To describe our algorithm, we first introduce some notation. Consider a collection of $J$ vectors $\{u_1, \dots, u_J\}$, each in $\mathbb{R}^2$. These $u_j$ are each endowed with a label $l_j \in \{1, \dots, L\}$ , indicating which category they are in – cases or controls. Also consider a second collection of $J'$ vectors $\{u'_1, \dots, u'_{J'}\}$, also in $\mathbb{R}^2$. The $u'_{j'}$ are also endowed with labels $l'_j \in \{1, \dots, L\}$. Given this notation, our goal will be to define an affine transformation $T : \mathbb{R}^2 \to \mathbb{R}^2$ which aligns the $\{u'_{j'}\}$ to the $\{u_J\}$ while correcting for the labels $\{l'_{j'}\}$ and $\{l_j\}$. This reduces the influence that the proportion of cases and controls can have on the transformation.

To begin, we'll first define the $k$-element vector $d(k; \{y_j\}, y') \in \mathbb{R}^k$, defined for a generic set $\{y_j\}$ and vector $y'$ as follows. We first find the $k$ nearest neighbors of $y'$ in the set $\{y_j\}$. Then we measure the distance between $y'$ and each of these $k$ nearest neighbors. Finally, we define $d(k; \{y_j\}, y')$ to be the $k$-element vector of these distances.

Using $d$ we can define the following symmetric error $E$ as follows:

$$E\left(k; \{y_j\}, \{y'_{j'}\}\right) = \frac{1}{k\left|\{y_{j'}\}\right|} \sum_{j'} \left\| d\left(k; \{y_j\}, y'_{j'}\right) \right\|^2 + \frac{1}{k\left|\{y_j\}\right|} \sum_{j} \left\| d\left(k; \{y'_{j'}\}, y_j\right) \right\|^2 ,$$

where $|\{\cdot\}|$ refers to the size of set $\{\cdot\}$, and each $\|d(k)\|^2$ is the sum of squares of the $k$ entries in that $d(k)$ (i.e., the squared frobenius-norm of $d(k)$).

Using $E$ and the labels $\{l_j\}$ and $\{l'_{j'}\}$, we can define a $T$-dependent error $F$ as follows:

$$F\left(T, k; \{x_j\}, \{l_j\}, \{x'_{j'}\}, \{l_{j'}\}\right) = \frac{1}{L} \sum_{l} E\left(k; \{x_j \,\middle|\, l_j = l\}, \{T \circ x'_{j'} \,\middle|\, l_{j'} = l\}\right)$$

Because the sets $\{x_j\}$, $\{l_j\}$, $\{x'_{j'}\}$ and $\{l_{j'}\}$ are fixed, we'll refer to $F$ below simply as $F(T, k)$.

Using this notion of error, we can define $T$ as follows:

**Initialize:** Set $T$ to be the identity transformation and set $k$ to be equal to $\max(J, J')$.

**Minimize:** Starting with the current estimate for $T$, use Nelder-Meade optimization to update $T$ by minimizing $F(T, k)$.

**Iterate:** Now reduce $k$ by one and return to the previous step.

**Terminate:** Stop when $k = 1$.

In practice we often observe that the final transformation $T$ is unchanged if the initial $k$ is sufficiently large. Here, we set $k = 32$. Similarly, the final transformation $T$ is often unchanged if $k$ is reduced by more than 1 each iteration.

#### ***Calculating label similarity***

To determine label similarity, we can take a particular point $y'_{j'}$ (i.e., an ADNI participant) with case-control label $l'_{j'}$, and compare it to the $k$ nearest neighbors in the set $\{y_j\}$ (i.e., UKB participants' data). The number of nearest neighbors with a matching label is denoted as $n(j, k)$. Then, we define $p(j, k) = n(j, k)/k$ to represent the fraction of nearest neighbors with a matching label. Then, we can define $\sigma(k; \{y_j\}, \{l_j\}, \{y'_{j'}\}, \{l'_{j'}\})$ as the average of $p(j, k)$ over $j$. That is, the average fraction of the $k$-nearest neighbors with the same label. We use linear interpolation to extend this to a continuous function in the interval $[0, |\{y_j\}|]$. Analogously, we can determine the similarity of UKB labels to ADNI labels, $\sigma(k';\ \{y'_{j'}\}, \{l'_{j'}\}, \{y_j\}, \{l_j\})$. Note that $\sigma(k';\ \{y'_{j'}\}, \{l'_{j'}\}, \{y_j\}, \{l_j\})$ is defined for all $k'$ in the interval $[0, |\{y'_{j'}\}|]$. Finally, for all $f \in [0,1]$, define we define the symmetric similarity as follows:

$$\tilde{\sigma}(f; \{y_j\}, \{l_j\}, \{y'_{j'}\}, \{l'_{j'}\}) = \frac{1}{2}\sigma(f|\{y_j\}|; \{y_j\}, \{l_j\}, \{y'_{j'}\}, \{l'_{j'}\}) + \frac{1}{2}\sigma(f|\{y'_j\}|; \{y'_{j'}\}, \{l'_{j'}\}, \{y_j\}, \{l_j\})$$

### Supplementary Tables

**Supplementary Table 1. Descriptive statistics of UK Biobank participants stratified by constellation and bicluster membership.**

| | Constellation 1 | | Constellation 2 | | Constellation 3 | |
|---|---|---|---|---|---|---|
| | Bicluster 1 cases | Non-bicluster | Bicluster 2 cases | Non-bicluster | Bicluster cases | Non-bicluster |
| **n** | 570 | 4423 | 190 | 2617 | - | 417 |
| **Sex, n male (%)** | 276 (48.4) | 2128 (48.1) | 92 (48.4) | 1297 (49.6) | - | 200 (48.0) |
| **Age, years (SD)** | 64.72 (4.32) | 64.75 (4.23) | 64.33 (4.65) | 64.75 (4.08) | - | 64.68 (4.46) |
| **Education, years (SD)** | 11.64 (4.85) | 11.94 (4.95) | 11.56 (4.78) | 12.09 (4.99) | - | 11.71 (5.02) |
| **AD dementia, n (%)** | 570 (100.0) | 1120 (25.3) | 190 (100.0) | 733 (28.0) | - | 126 (30.2) |
| **APOE-e4 alleles, n (%)** | | | | | | |
| **0** | 212 (37.2) | 2852 (64.5) | 79 (41.6) | 1676 (64.0) | - | 263 (63.1) |
| **1** | 284 (49.8) | 1353 (30.6) | 78 (41.1) | 799 (30.5) | - | 133 (31.9) |
| **2** | 74 (13.0) | 218 (4.9) | 33 (17.4) | 142 (5.4) | - | 21 (5.0) |

**Supplementary Table 2. Genotypes of *MAPT* haplotype H1/H2-tagging SNP rs8070723 across constellations.** The SNP rs8070723 may be used to determine haplotype carriership, with the A allele associated with haplotype H1 and the G allele associated with haplotype H2. There was a strong, but not perfect, correspondence between genotype and constellation.

| | **Constellation 1** | **Constellation 2** | **Constellation 3** |
|---|---|---|---|
| **AA** | 4985 | 1 | 2 |
| **AG** | 8 | 2802 | 6 |
| **GG** | 0 | 4 | 409 |

### Supplementary Figures

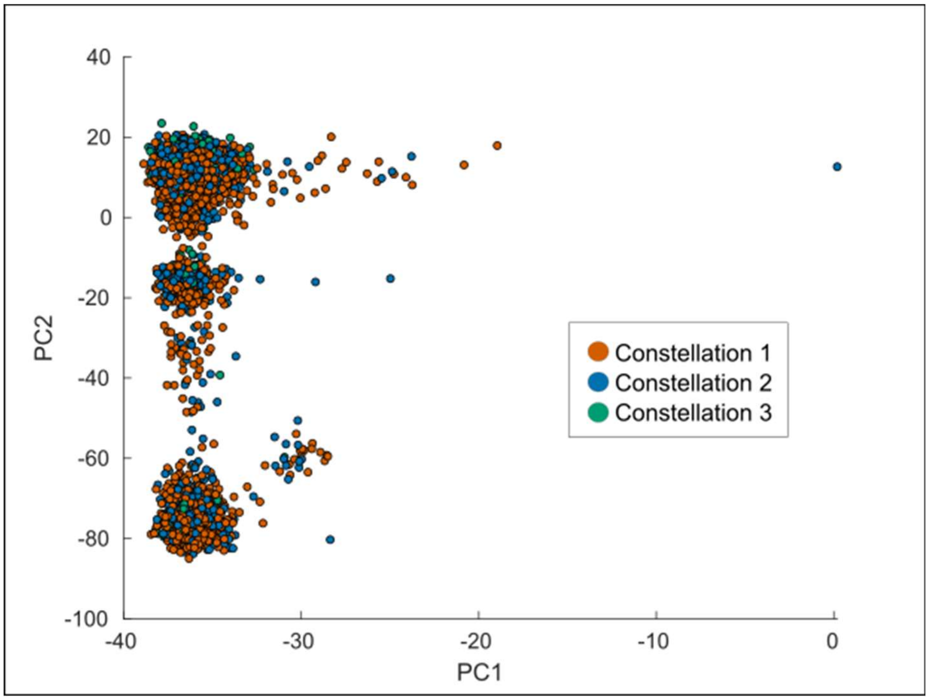


**Supplementary Figure 1. Principal components analysis of a random subset of variants.** Principal components analysis (PCA) was applied to allele combinations of UK Biobank cases and controls from a random selection of variants. The number of variants was identical to the number included in the analysis of Alzheimer's-associated variants (i.e., p<0.05 in the Kunkle et al. (2019) Alzheimer's GWAS). The scatter plots display participant loadings on the first two principal components (PC1 and PC2) and colored by constellation labels defined from the analysis of at the p<0.05 threshold. Participants from all three constellations are highly mixed and mirrors the structure found in the analysis that included all (i.e., p<1.0) variants. This indicates that the constellation structure is not simply a function of the number of variants included in the analysis but emerges among disease-relevant variants.

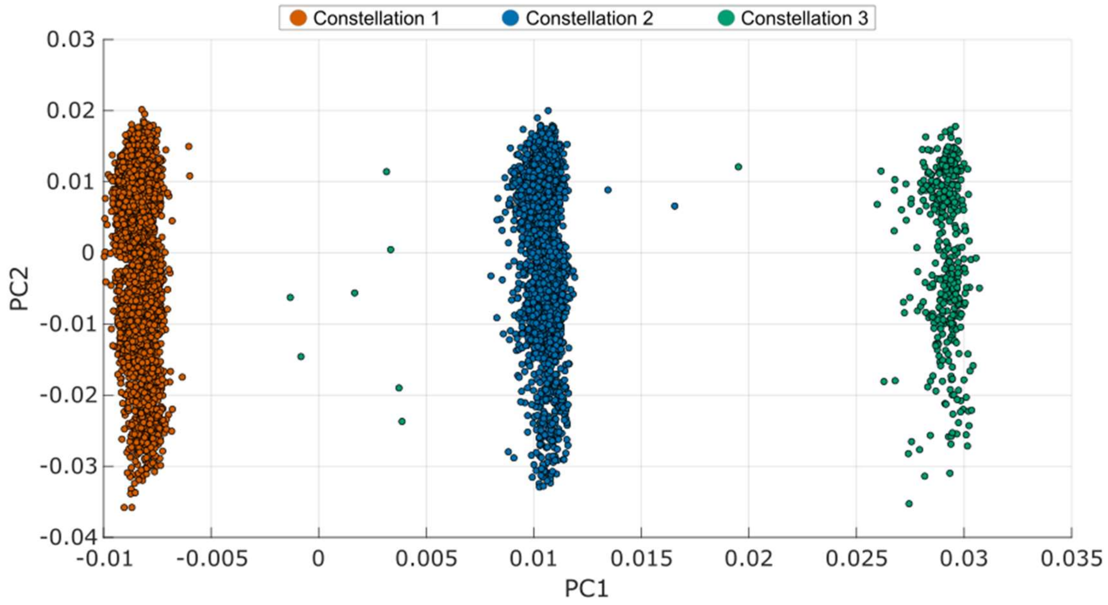


**Supplementary Figure 2. Principal components analysis of additively-coded data.** Principal component analysis was applied to additively-coded (i.e. dosage-coded) data from UK Biobank cases and controls restricted to variants with a p-value<0.05 in the Kunkle et al. Alzheimer's GWAS [79]. The scatter plot displays participant loading on the first two principal components (PC1 and PC2) and are colored by constellation groups defined by the PCA of allele-coded data used in the primary analysis and shown in Figures 1 and 5 in the main text. Comparison of these plots indicates that the distinct clustering is not a by-product of the encoding scheme used.

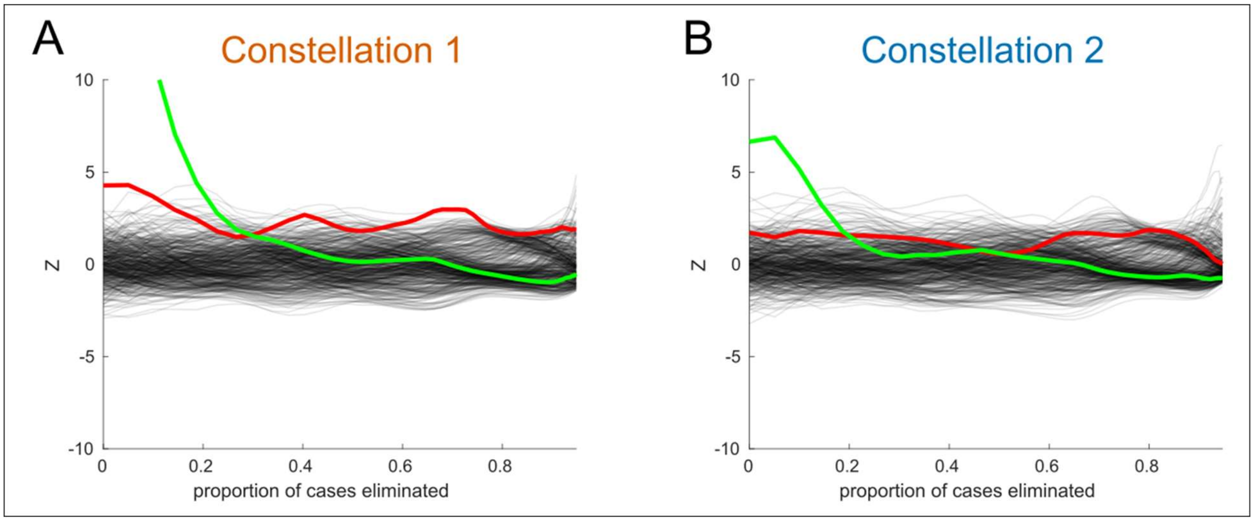


**Supplementary Figure 3. Traces of search for primary and secondary biclusters.** The disease-related signal-strength associated with the remaining UKB Alzheimer's cases relative to controls is plotted on the y-axis for A) bicluster 1 (founding in constellation 1) and B) bicluster 2 (found in constellation 2). At each iteration, allele combinations and cases that contribute least to this difference are removed. The proportion of remaining cases is shown on the x-axis. The red trace represents the original data and black traces represent label-shuffled data, corresponding to a null distribution. After identifying the primary bicluster, it is removed by scrambling the entries of the submatrix associated with bicluster (i.e., entries corresponding to the participants and allele-combinations that were retained in the bicluster) and the search algorithm is run again. The green trace represents this partially scrambled data. Constellation 3 is not shown because no significant primary bicluster was found.

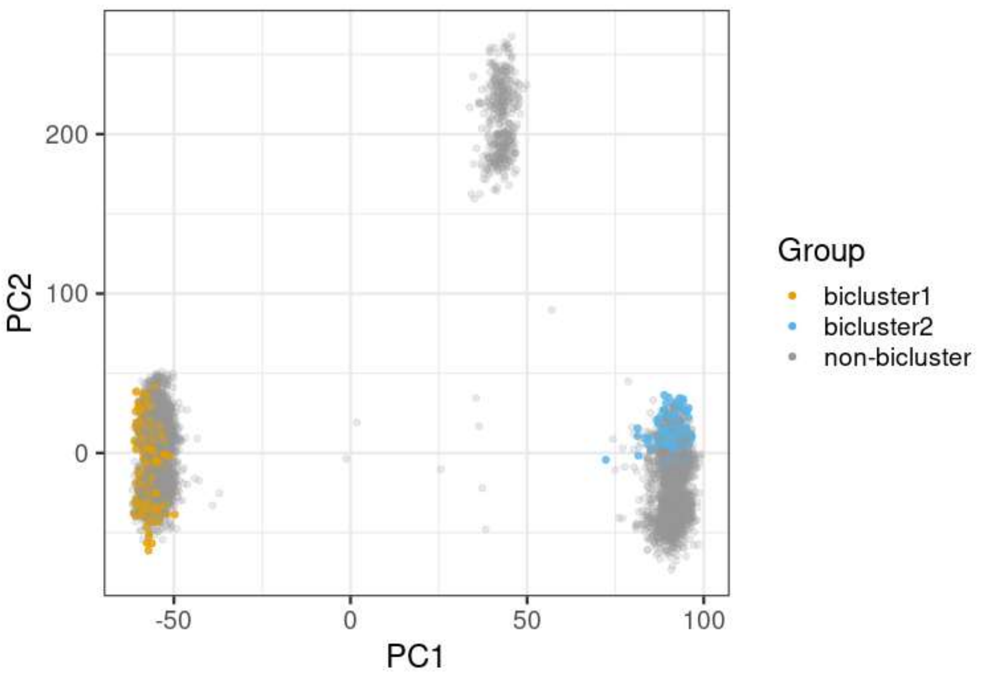


**Supplementary Figure 4. Principal components analysis of UKB cases and controls colored by bicluster membership.** Principal component analysis was applied to allele combinations of UK Biobank cases and controls restricted to variants with a p-value<0.05 in the Kunkle et al. Alzheimer's GWAS (12). The first two PCs are plotted as in Figure 1 from the main text, but individuals are now colored according to membership in a bicluster.

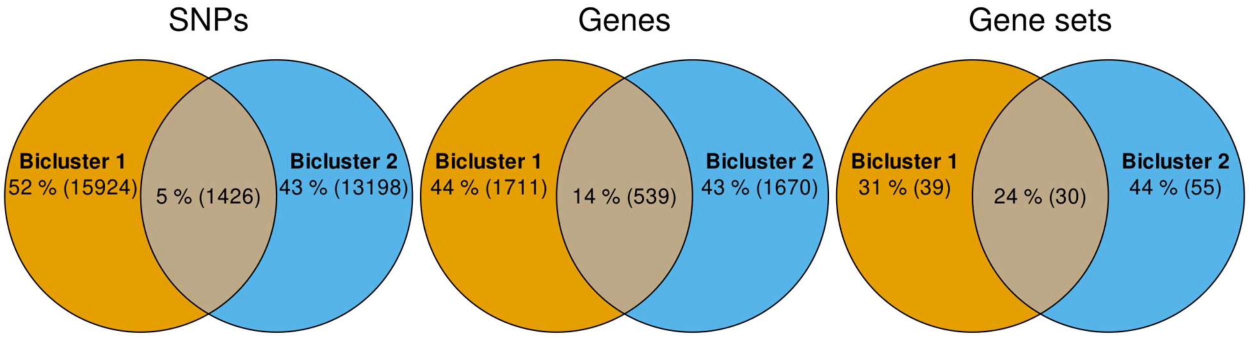


**Supplementary Figure 5. Overlap in gene sets, genes, and SNPs between biclusters.** Panels A-C show the number of unique and shared elements between disease-specific biclusters. Separate GWAS compared bicluster cases to controls in the same constellation (e.g., bicluster 1 cases versus controls also belonging to constellation 1). A) Overlap in SNPs associated with each constellation or bicluster. B) Overlap in the corresponding genes to which SNPs associated with each bicluster were able to be mapped. C) Overlap in gene sets significantly enriched in each bicluster based on over-representation analysis of gene lists. In general, the amount of overlap was greater at higher levels of aggregation (e.g., there was more overlap of gene sets than genes or SNPs).

### REFERENCES


[1] Rangan AV, McGrouther CC, Kelsoe J, Schork N, Stahl E, Zhu Q, Krishnan A, Yao V, Troyanskaya O, Bilaloglu S, Raghavan P, Bergen S, Jureus A, Landen M, Bipolar Disorders Working Group of the Psychiatric Genomics C (2018) A loop-counting method for covariate-corrected low-rank biclustering of gene-expression and genome-wide association study data. *PLoS Comput Biol* **14**, e1006105.

[2] Zhou H, Lin W, Labra SR, Lipton SA, Schork NJ, Rangan AV (2022) Detecting boolean asymmetric relationships with a loop counting technique and its implications for analyzing heterogeneity within gene expression datasets. *bioRxiv*, 2022.2008.2004.502792.

[3] Febrero M, Galeano P, González-Manteiga W (2007) Outlier detection in functional data by depth measures, with application to identify abnormal NOx levels. *Environmetrics* **19**, 331-345.
